# Occupational differences in the prevalence and severity of long-COVID: Analysis of the ONS Coronavirus (COVID-19) Infection Survey

**DOI:** 10.1101/2023.03.24.23287666

**Authors:** Theocharis Kromydas, Evangelia Demou, Rhiannon Edge, Matthew Gittins, S Vittal Katikireddi, Neil Pearce, Martie van Tongeren, Jack Wilkinson, Sarah Rhodes

## Abstract

**Objectives:** To establish whether prevalence and severity of long-COVID symptoms vary by industry and occupation.

**Methods:** We utilised ONS Coronavirus Infection Survey (CIS) data (February 2021-April 2022) of working-age participants (16-65 years). Exposures were industrial sector, occupation and major Standard Occupational Classification (SOC) group. Outcomes were self-reported: (1) long-COVID symptoms; and (2) reduced function due to long-COVID. Binary (outcome 1) and ordered (outcome 2) logistic regression were used to estimate odds ratios (OR) and prevalence (marginal means) for all exposures.

**Results:** Public facing industries, including teaching and education, social care, healthcare, civil service, retail and transport industries and occupations had highest odds ratios for long-COVID. By major SOC group, those in caring, leisure and other services (OR 1.44, CIs: 1.38-1.52) had substantially elevated odds than average. For almost all exposures, the pattern of odds ratios for long-COVID symptoms followed that for SARS-CoV-2 infections, except for professional occupations (OR<1 for infection; OR>1 for long-COVID). The probability of reporting long-COVID for industry ranged from 7.7% (financial services) to 11.6% (teaching and education); whereas the prevalence of reduced function by ‘a lot’ ranged from 17.1% (arts, entertainment and recreation) to 22-23% (teaching and education and armed forces) and to 27% (those not working).

**Conclusions:** The risk and prevalence of long-COVID differs across industries and occupations. Generally, it appears that likelihood of developing long-COVID symptoms follows likelihood of SARS-CoV-2 infection, except for professional occupations. These findings highlight sectors and occupations where further research is needed to understand the occupational factors resulting in long-COVID.

**Key messages:** 

**What is already known on this topic:**

- SARS-CoV-2 infection and COVID-19 mortality in the UK varied by occupational group; yet it is not known if any occupational groups are more susceptible to long-COVID than others.

**What this study adds:**

- This is the first study to examine how prevalence of long-COVID and its impacts on functional capacity differ by industrial sector and occupational groups.
- Prevalence of self-reported long-COVID increased with time across all exposure groups and mostly followed SARS-CoV-2 infection trends; with the exception of Professional occupations that demonstrated notable differences in the direction of odds of long-covid when compared to odds of SARS-CoV-2 infection.
- Those working in Teaching and education, and social care industries showed the highest likelihood of having long-COVID symptoms. The exact same pattern was observed when analysis was performed using occupational groups. When we used SOC groups the likelihood was higher in Caring, leisure and other services.

**How this study might affect research, practice or policy:**

- The findings contribute to the evidence base that long-COVID differences occur across industries and occupations, provides insights for employees, employers, occupational and healthcare for the industries and occupations that may need additional support for return- to-work policies and highlights sectors and occupations where further research is needed to understand the mechanisms resulting in long-COVID and how occupational factors influence the risk of developing long-COVID or interact with long-COVID to increase the impact on activities.

## Introduction

In the UK, the risk of SARS-CoV-2 infection and COVID-19 mortality has varied by occupational group, although most differences appear to have declined over the duration of the pandemic (1-5). It remains unclear however, whether or not some occupational groups are more susceptible to long-; and if any differences reflect, or are addition to, differential risks in SARS-CoV-2 infection (6). The highest risks of long-COVID are reported amongst workers in education, social care and healthcare sectors (7); all sectors with elevated risks of SARS-CoV-2 infection during the pandemic (8).

Being out of work is associated with poor health (9-11) and the risk unemployment increases with length of sick leave (12). Recent studies have demonstrated the burden of long-COVID, where even after months, many patients report persisting symptoms and have not returned to previous levels of work (13 14). A Danish nationwide registry study demonstrated that of those hospitalised, being female, older age, and having a comorbidity were associated with a lower chance of returning to work (15). Davies **et al**, showed that almost half of the study respondents suffering with long-COVID required a reduced work schedule compared to pre-illness (13). A Swedish national cohort study showed that 13% were on sick leave due to long-COVID from March-August 2020 and 9% were on sick leave for at least four months (16). Those on sick leave due to long-COVID were older, predominantly men, spent more time on sick leave prior to COVID-19, and were more likely to have received inpatient care (16). While we understand some of the factors that predict long-COVID and work ability, little is known about how the prevalence of long-COVID and associated functional capacity differs by occupation and the impact of occupational factors on these patterns.

Long-COVID has a disproportionate impact on groups already disadvantaged in terms of work and health. Returning to work is part of rehabilitation from illness and is important to those recovering from long-COVID (12). Many people with significant illness or disability work effectively if they are provided with suitable support in the workplace (12). Better understanding of the prevalence and severity of long-COVID in different occupational groups will inform the need for rehabilitation, workplace adjustment measures and support (17) and provide evidence about whether or not long-COVID should be considered an occupational disease (7). It may also identify workplaces that need additional support measures to reduce risks of transitioning to long-COVID after SARS-CoV-2 infection.

Our study aims to establish whether the prevalence and severity of long-COVID varied by occupation and if any differences are in excess of differences in the risk of SARS-CoV-2 infection.

## METHODS AND DATA

### Study design

We used data from the ONS Coronavirus (COVID-19) Infection Survey (CIS) from Feb 2021 (when long-Covid questions were first included in CIS) until the end of April 2022. This longitudinal study began in April 2020, used random sampling and aimed to be representative of the UK population. Repeated PCR testing for all participants was carried out during weekly home visits for the first month of entry to the survey, and then monthly until 30^th^ April 2022 when testing samples were sent and returned by post and survey questions were asked on-line or over the phone (18). CIS uses 4 different variables to capture employment status. Participants were coded as employed or not working (Employment status coding section).

Analyses were restricted to participants aged ≥16 to ≤65 years old on the day of their first home visit and to those who turned 16 during the study period and had answered long-COVID questions at least once.

We used the first available observation per occupational exposure and other covariates, assuming no change across time. In sensitivity analyses, we estimated panel models to allow for variation across time.

### Exposure groups

We used three different industry and occupational groupings. Industry was classified based on the ONS sector groupings used within CIS (19). Based on expert judgement and consensus among team members we derived a bespoke set of occupational groupings using 4-digit standard occupational codes (SOC 2010) (Table S1) (20). Groups used in previous studies (2-4 21) focussed on key worker/essential worker groups (i.e., those with highest exposure at the start of the pandemic) and therefore new groupings were needed to capture all occupations and exposures while ensuring adequate group size, as the pandemic progressed. The third exposure category was by major SOC group.

### Analytical sample and ascertainment of outcome

Primary outcome measures were self-reported (i) cases of long-COVID and (ii) reduced function (i.e., severity) due to long-COVID among those reporting long-COVID, using the following CIS questions:

i. self-reported long-COVID: *“Would you describe yourself as having ‘long-COVID’, that is, you are still experiencing symptoms more than 4 weeks after you first had COVID-19, that are not explained by something else?”*. For our primary analysis we reported the n (%) of people who self-reported having long-COVID in at least one survey, irrespective of any COVID-19 test result or previous infection reported.
ii. self-reported reduced function (severity) of long-COVID: *‘How much did long-COVID reduce your ability to carry out daily activities?’* Respondents could answer 0: not at all, 1: Yes, a little, 2: Yes, a lot. This analytical sample was restricted to those self-reporting long-covid symptoms.

### Statistical analyses

Analysis was completed for the two outcome variables using all three exposure groups, irrespective of whether they have tested or reported being previously infected by SARS-CoV-2. For comparison, we also performed analyses using the outcome ‘SARS-CoV-2 infection (self-reported or positive PCR test).

Sample characteristics were summarised using frequencies and proportions. Descriptive statistics were used to show the change in prevalence of self-reported long-COVID over time. Binary (outcome 1: long-COVID symptoms) and ordered (outcome 2: reduced function due to long-COVID) logistic regression models with robust standard errors to account for heteroskedasticity were used. For the binary logistic regression analyses, data were collapsed by person using the first reported values for exposure and covariates and taking ‘at least one reported episode of long-COVID’ as the outcome. For the ordered logistic regression, the highest reported value for the outcome variable (i.e., severity of long-COVID) was used per person. We used effects coding where ORs are contrasted to each group mean compared to the grand mean showing the likelihood of having long-COVID in each group. To assess for potential confounding or mediating differences we estimated unadjusted and fully adjusted models for all the covariates (age, sex, IMD, rural or urban location, household size, region, pre-existing health conditions) (2).

Weighting was not used as the CIS derived weights are cross sectional. Testing for patterns of missingness in the outcome variable did not show any differences (Table S2). Participants with no measures in our outcome, exposure and covariate variables were excluded (Figure 1; Table S2). We included the non-working group for comparison purposes.

**Figure 1.**
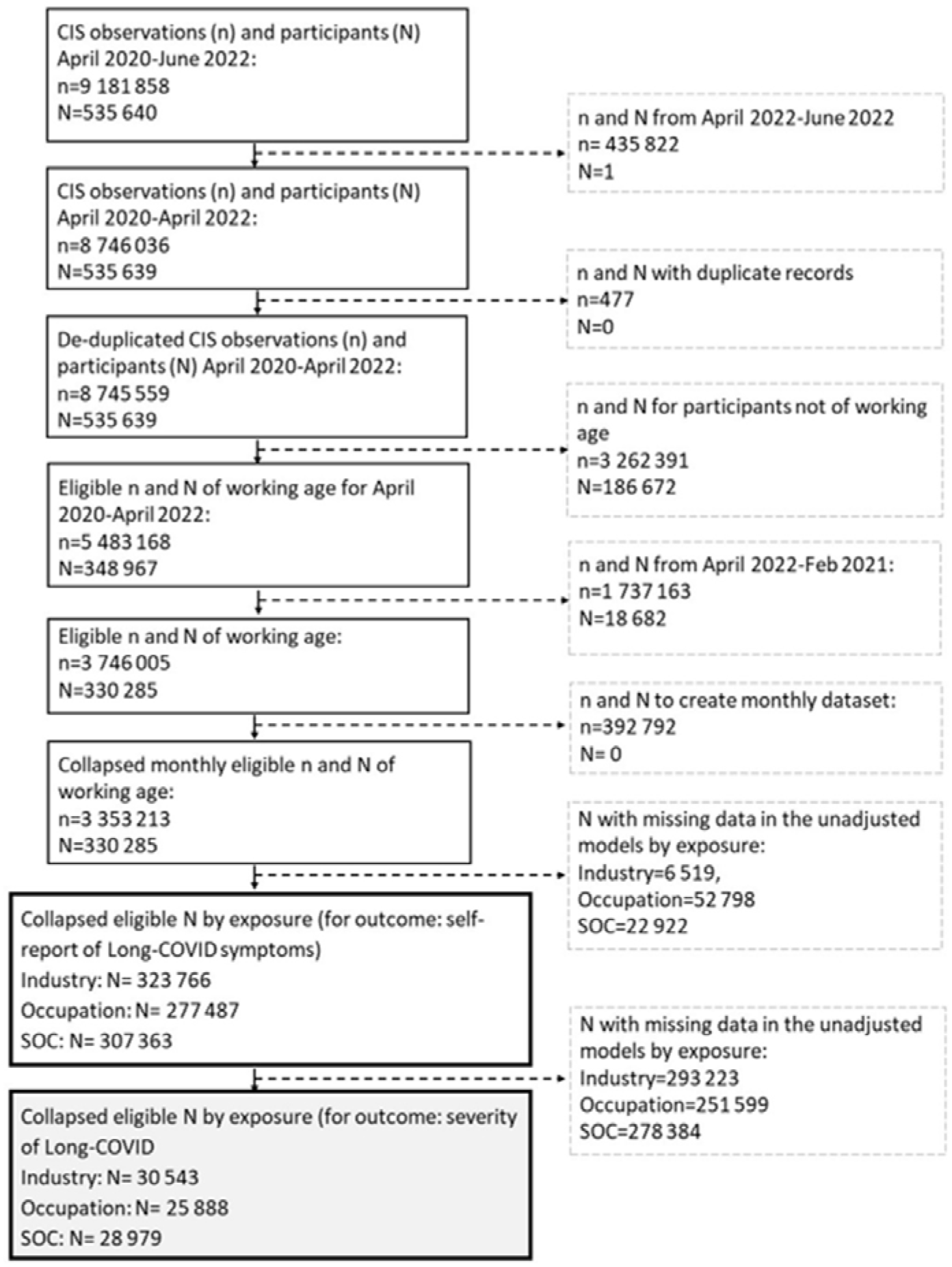
Flow chart of Covid Infection Survey (CIS) participants. N= number of participants, n=number of observations

#### Sensitivity Analyses

Sensitivity analyses based on our main analytical methods was conducted using two additional analytical samples: (i) those with a positive PCR test; and (ii) those with a positive PCR test and/or self-reported infection before entering the CIS. Furthermore, using the specifications of our main sample, multiple participant visits and records were merged where needed to create a dataset with one observation per month per person to allow variation across time (Figure 1). We used multilevel mixed-methods generalised linear models, with a binomial link function for the outcome of having long-COVID symptoms; and a multilevel mixed-effects ordered logistic regression for the outcome of reduced function due to long-COVID and compare with the estimations of our analytical model.

All analyses were calculated using STATA 17 (22).

## RESULTS

### Cohort characteristics

Our final analytical samples included 323 766 individuals by industry, 277 487 individuals by occupational group and 307 363 individuals by major SOC group (Figure 1). Table 1 illustrates descriptive statistics by exposure group. In all cases, the non-working group had approximately twice as high prevalence of underlying health conditions (Table S3) and a higher prevalence of being affected ‘a lot’ by long-COVID. (Tables S4b, S5b, S6b) compared to the working group.

**Table 1.** Descriptive statistics of all covariates used for adjustment by exposure and self-reported long-covid case-Number and percentages of individuals

| Exposure groups |  | Industry (SIC) |  |  |  |  |  | Occupational groups |  |  |  |  |  | Major occupational groups (SOC) |  |  |  |  |  |
| --- | --- | --- | --- | --- | --- | --- | --- | --- | --- | --- | --- | --- | --- | --- | --- | --- | --- | --- | --- |
| Outcome | Self-reported long-COVID | No | Count<br>Yes | Total | No | %<br>Yes | Total | No | Count<br>Yes | Total | No | %<br>Yes | Total | No | Count<br>Yes | Total | No | %<br>Yes | Total |
| Age bands | 15-19 | 21 083 | 1 440 | 22 523 | 7.2 | 4.7 | 7.0 | 20 771 | 1 415 | 22 186 | 8.3 | 5.5 | 8.0 | 20 911 | 1 430 | 22 341 | 7.5 | 4.9 | 7.3 |
|  | 20-24 | 15 820 | 1 140 | 16 960 | 5.4 | 3.7 | 5.2 | 14 149 | 998 | 15 147 | 5.6 | 3.9 | 5.5 | 15 193 | 1 089 | 16 282 | 5.5 | 3.8 | 5.3 |
|  | 25-29 | 20 191 | 1 516 | 21 707 | 6.9 | 5.0 | 6.7 | 16 607 | 1 234 | 17 841 | 6.6 | 4.8 | 6.4 | 18 864 | 1 432 | 20 296 | 6.8 | 4.9 | 6.6 |
|  | 30-34 | 24 958 | 2 168 | 27 126 | 8.5 | 7.1 | 8.4 | 20 492 | 1 780 | 22 272 | 8.1 | 6.9 | 8.0 | 23 390 | 2 045 | 25 435 | 8.4 | 7.1 | 8.3 |
|  | 35-39 | 27 647 | 2 911 | 30 558 | 9.4 | 9.5 | 9.4 | 22 596 | 2 348 | 24 944 | 9.0 | 9.1 | 9.0 | 25 890 | 2 715 | 28 605 | 9.3 | 9.4 | 9.3 |
|  | 40-44 | 29 224 | 3 766 | 32 990 | 10.0 | 12.3 | 10.2 | 23 787 | 3 065 | 26 852 | 9.5 | 11.8 | 9.7 | 27 315 | 3 520 | 30 835 | 9.8 | 12.2 | 10.0 |
|  | 45-49 | 30 989 | 4 032 | 35 021 | 10.6 | 13.2 | 10.8 | 25 318 | 3 323 | 28 641 | 10.1 | 12.8 | 10.3 | 28 983 | 3 807 | 32 790 | 10.4 | 13.1 | 10.7 |
|  | 50-54 | 34 809 | 4 550 | 39 359 | 11.9 | 14.9 | 12.2 | 28 866 | 3 752 | 32 618 | 11.5 | 14.5 | 11.8 | 32 671 | 4 270 | 36 941 | 11.7 | 14.7 | 12.0 |
|  | 55-59 | 38 180 | 4 353 | 42 533 | 13.0 | 14.3 | 13.1 | 32 773 | 3 744 | 36 517 | 13.0 | 14.5 | 13.2 | 36 252 | 4 146 | 40 398 | 13.0 | 14.3 | 13.1 |
|  | 60-64 | 39 819 | 3 819 | 43 638 | 13.6 | 12.5 | 13.5 | 36 261 | 3 435 | 39 696 | 14.4 | 13.3 | 14.3 | 38 585 | 3 694 | 42 279 | 13.9 | 12.8 | 13.8 |
|  | 65-69 | 10 503 | 848 | 11 351 | 3.6 | 2.8 | 3.5 | 9 979 | 794 | 10 773 | 4.0 | 3.1 | 3.9 | 10 329 | 832 | 11 161 | 3.7 | 2.9 | 3.6 |
| Sex | Male | 135 306 | 11 759 | 147 065 | 46.1 | 38.5 | 45.4 | 114 942 | 9 826 | 124 768 | 45.7 | 38.0 | 45.0 | 127 779 | 11 075 | 138 854 | 45.9 | 38.2 | 45.2 |
|  | Female | 157 917 | 18 784 | 176 701 | 53.9 | 61.5 | 54.6 | 136 657 | 16 062 | 152 719 | 54.3 | 62.0 | 55.0 | 150 604 | 17 905 | 168 509 | 54.1 | 61.8 | 54.8 |
| Deprivation | 1st Quartile | 44 738 | 5 668 | 50 406 | 15.3 | 18.6 | 15.6 | 39 010 | 4 908 | 43 918 | 15.5 | 19.0 | 15.8 | 42 574 | 5 393 | 47 967 | 15.3 | 18.6 | 15.6 |
|  | 2nd Quartile | 68 677 | 7 360 | 76 037 | 23.4 | 24.1 | 23.5 | 58 714 | 6 292 | 65 006 | 23.3 | 24.3 | 23.4 | 65 160 | 7 007 | 72 167 | 23.4 | 24.2 | 23.5 |
|  | 3rd Quartile | 83 677 | 8 398 | 92 075 | 28.5 | 27.5 | 28.4 | 71 560 | 7 095 | 78 655 | 28.4 | 27.4 | 28.4 | 79 472 | 7 958 | 87 430 | 28.6 | 27.5 | 28.5 |
|  | 4th Quartile | 96 131 | 9 117 | 105 248 | 32.8 | 29.9 | 32.5 | 82 315 | 7 593 | 89 908 | 32.7 | 29.3 | 32.4 | 91 177 | 8 622 | 99 799 | 32.8 | 29.8 | 32.5 |
| Government office regions | North East | 10 033 | 1 349 | 11 382 | 3.4 | 4.4 | 3.5 | 8 563 | 1 124 | 9 687 | 3.4 | 4.3 | 3.5 | 9 540 | 1 279 | 10 819 | 3.4 | 4.4 | 3.5 |
|  | North West | 32 049 | 3 914 | 35 963 | 10.9 | 12.8 | 11.1 | 27 026 | 3 241 | 30 267 | 10.7 | 12.5 | 10.9 | 30 258 | 3 678 | 33 936 | 10.9 | 12.7 | 11.0 |
|  | Yorkshire & the Humber | 23 074 | 2 692 | 25 766 | 7.9 | 8.8 | 8.0 | 19 772 | 2 264 | 22 036 | 7.9 | 8.8 | 7.9 | 21 969 | 2 571 | 24 540 | 7.9 | 8.9 | 8.0 |
|  | East Midlands | 18 474 | 2 032 | 20 506 | 6.3 | 6.7 | 6.3 | 15 832 | 1 728 | 17 560 | 6.3 | 6.7 | 6.3 | 17 544 | 1 938 | 19 482 | 6.3 | 6.7 | 6.3 |
|  | West Midlands | 21 127 | 2 430 | 23 557 | 7.2 | 8.0 | 7.3 | 18 163 | 2 033 | 20 196 | 7.2 | 7.9 | 7.3 | 20 160 | 2 320 | 22 480 | 7.2 | 8.0 | 7.3 |
|  | East of England | 26 605 | 2 704 | 29 309 | 9.1 | 8.9 | 9.1 | 22 254 | 2 230 | 24 484 | 8.9 | 8.6 | 8.8 | 25 298 | 2 577 | 27 875 | 9.1 | 8.9 | 9.1 |
|  | London | 56 565 | 5 190 | 61 755 | 19.3 | 17.0 | 19.1 | 46 784 | 4 276 | 51 060 | 18.6 | 16.5 | 18.4 | 52 959 | 4 886 | 57 845 | 19.0 | 16.9 | 18.8 |
|  | South East | 36 248 | 3 477 | 39 725 | 12.4 | 11.4 | 12.3 | 31 041 | 2 919 | 33 960 | 12.3 | 11.3 | 12.2 | 34 384 | 3 282 | 37 666 | 12.4 | 11.3 | 12.3 |
|  | South West | 22 150 | 2 147 | 24 297 | 7.6 | 7.0 | 7.5 | 19 331 | 1 861 | 21 192 | 7.7 | 7.2 | 7.6 | 21 195 | 2 021 | 23 216 | 7.6 | 7.0 | 7.6 |
|  | Northern Ireland | 8 378 | 838 | 9 216 | 2.9 | 2.7 | 2.9 | 7 431 | 738 | 8 169 | 3.0 | 2.9 | 2.9 | 7 851 | 785 | 8 636 | 2.8 | 2.7 | 2.8 |
|  | Scotland | 24 138 | 2 266 | 26 404 | 8.2 | 7.4 | 8.2 | 22 263 | 2 110 | 24 373 | 8.9 | 8.2 | 8.8 | 23 428 | 2 205 | 25 633 | 8.4 | 7.6 | 8.3 |
|  | Wales | 14 382 | 1 504 | 15 886 | 4.9 | 4.9 | 4.9 | 13 139 | 1 364 | 14 503 | 5.2 | 5.3 | 5.2 | 13 797 | 1 438 | 15 235 | 5.0 | 5.0 | 5.0 |
| Urban or Rural | Major urban | 110 000 | 11 547 | 121 547 | 37.5 | 37.8 | 37.5 | 92 337 | 9 624 | 101 961 | 36.7 | 37.2 | 36.7 | 103 478 | 10 918 | 114 396 | 37.2 | 37.7 | 37.2 |
|  | Urban city or town | 122 942 | 13 315 | 136 257 | 41.9 | 43.6 | 42.1 | 106 614 | 11 333 | 117 947 | 42.4 | 43.8 | 42.5 | 117 243 | 12 617 | 129 860 | 42.1 | 43.5 | 42.3 |
|  | Rural town | 29 107 | 2 890 | 31 997 | 9.9 | 9.5 | 9.9 | 25 176 | 2 494 | 27 670 | 10.0 | 9.6 | 10.0 | 27 806 | 2 790 | 30 596 | 10.0 | 9.6 | 10.0 |
|  | Rural village | 31 174 | 2 791 | 33 965 | 10.6 | 9.1 | 10.5 | 27 472 | 2 437 | 29 909 | 10.9 | 9.4 | 10.8 | 29 856 | 2 655 | 32 511 | 10.7 | 9.2 | 10.6 |
| Household Size | One | 38 251 | 4 202 | 42 453 | 13.1 | 13.8 | 13.1 | 32 671 | 3 562 | 36 233 | 13.0 | 13.8 | 13.1 | 36 288 | 3 975 | 40 263 | 13.0 | 13.7 | 13.1 |
|  | Two | 109 005 | 10 377 | 119 382 | 37.2 | 34.0 | 36.9 | 93 306 | 8 859 | 102 165 | 37.1 | 34.2 | 36.8 | 103 553 | 9 881 | 113 434 | 37.2 | 34.1 | 36.9 |
|  | Three | 59 258 | 6 298 | 65 556 | 20.2 | 20.6 | 20.3 | 50 670 | 5 290 | 55 960 | 20.1 | 20.4 | 20.2 | 56 126 | 5 972 | 62 098 | 20.2 | 20.6 | 20.2 |
|  | Four | 59 824 | 6 646 | 66 470 | 20.4 | 21.8 | 20.5 | 51 287 | 5 627 | 56 914 | 20.4 | 21.7 | 20.5 | 56 783 | 6 303 | 63 086 | 20.4 | 21.8 | 20.5 |
|  | Five plus | 26 885 | 3 020 | 29 905 | 9.2 | 9.9 | 9.2 | 23 665 | 2 550 | 26 215 | 9.4 | 9.9 | 9.5 | 25 633 | 2 849 | 28 482 | 9.2 | 9.8 | 9.3 |
| Health Condition | No | 248 054 | 23 727 | 271 781 | 84.6 | 77.7 | 83.9 | 210 591 | 19 808 | 230 399 | 83.7 | 76.5 | 83.0 | 234 681 | 22 422 | 257 103 | 84.3 | 77.4 | 83.7 |
|  | Yes | 45 169 | 6 816 | 51 985 | 15.4 | 22.3 | 16.1 | 41 008 | 6 080 | 47 088 | 16.3 | 23.5 | 17.0 | 43 702 | 6 558 | 50 260 | 15.7 | 22.6 | 16.4 |
| **Total** |  | **293 223** | **30 543** | **323 766** | **90.6** | **9.4** | **100.0** | **251 599** | **25 888** | **277 487** | **90.7** | **9.3** | **100.0** | **278 383** | **28 980** | **307 363** | **90.6** | **9.4** | **100.0** |

Monthly prevalence of self-reported long-COVID and SARS-CoV-2 infection across time and across all three occupational exposures, demonstrates an increase of the prevalence of self-reported long-COVID with time, following similar trends in SARS-CoV-2 infection (Figure 2). Prevalence of long-COVID by industry ranges from ∼2% to almost 6% in April 2022 for social care and education. The healthcare sector also shows relatively high prevalence that increases post-January 2022. Similar patterns in long-COVID prevalence are observed by occupation and major SOC group. Education and social care and hospitality occupations demonstrate the highest prevalence (>4% from Jan 2022). Caring, leisure and other service occupations (SOC group) exhibit the highest prevalence, followed by sales and customer service occupations.

**Figure 2:**
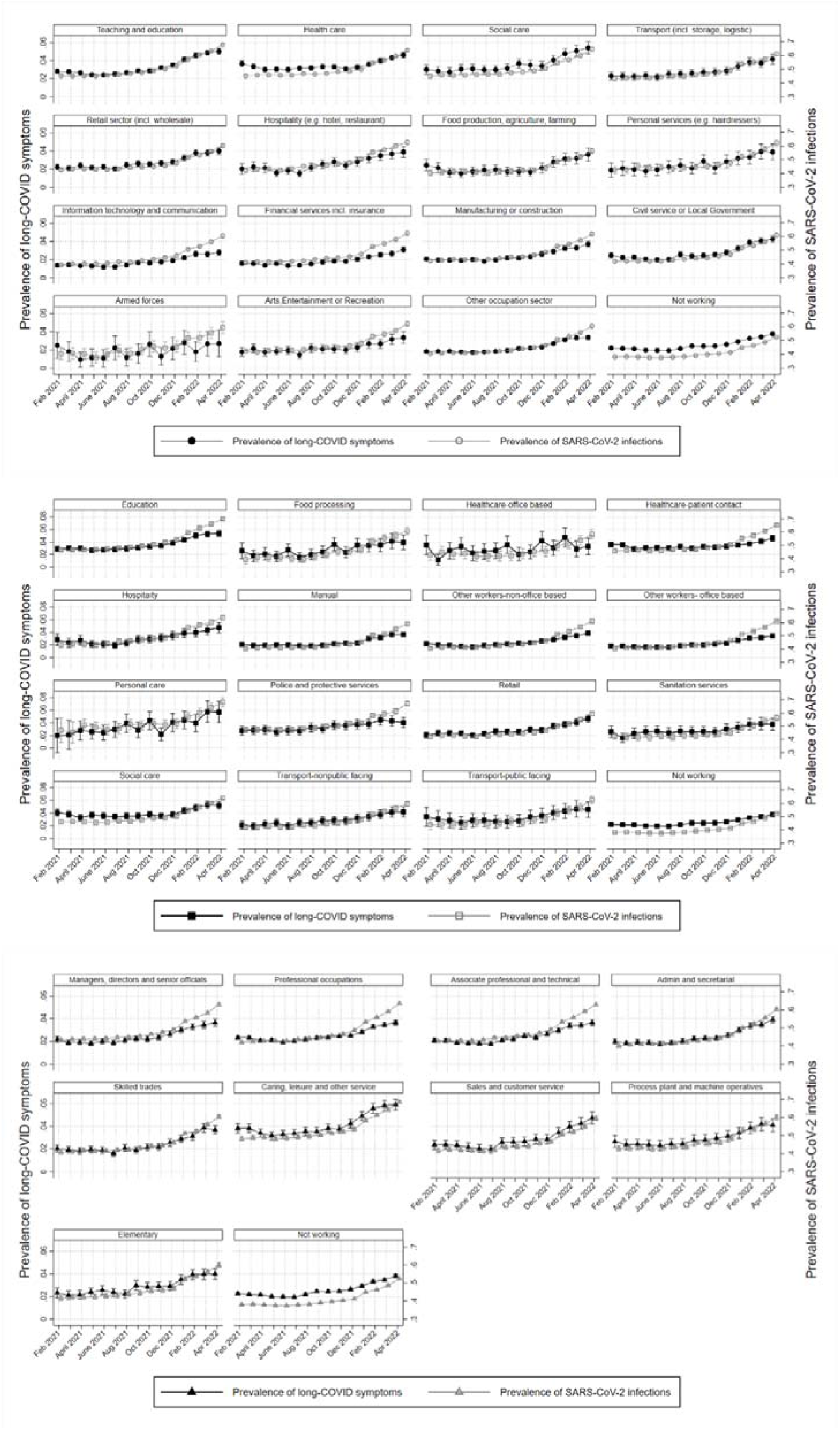
Monthly prevalence of self-reported long-COVID symptoms (Primary y-axis; left) and SARS-CoV-2 infections (secondary y-axis; right) for all workers aged 16-65 for (a) industry; (b) occupational groups: and (c) SOC major groups

## Relative risk (ORs) and prevalence (marginal means) of long-COVID

In the unadjusted model (Table S7), Teaching and education (OR 1.41; CIs: 1.36-1.46), Social care (OR 1.38, CIs: 1.28-1.48), Healthcare (1.19; CIs: 1.14-1.23), Civil service or local government (1.12; CIs: 1.06-1.17), retail (1.08; CIs: 1.08; 1.03-1.14) and transport (1.08; CIs: 1.00-1.16) demonstrated increased likelihood of reporting long-COVID symptoms compared to the average working-age population. All other sectors (including the non-working group) showed odds ratios (ORs) less than one or with very wide CIs, with the lowest ORs being in the information technology and communication sector(IT) (0.75; CIs: 0.71-0.79). In the fully adjusted model (Table S7, Figure 3a), ORs for most industries were attenuated and the pattern remained the same with Teaching and education exhibiting the highest odds (1.27; CIs: 1.23-1.31). Exceptions to confounding attenuation were primarily seen for the transport industry (1.12; CIs: 1.04-1.21) and hospitality and manufacturing, albeit with wide confidence intervals. Prevalence of long-COVID (marginal means-predicted probability of having at least one episode of long-COVID symptoms across all groups) ranged from 7.8% (IT, financial services and armed forces) to 11.1% (social care) and 11.6% for teaching and education (Table S9, Figure S1a).

**Figure 3.**
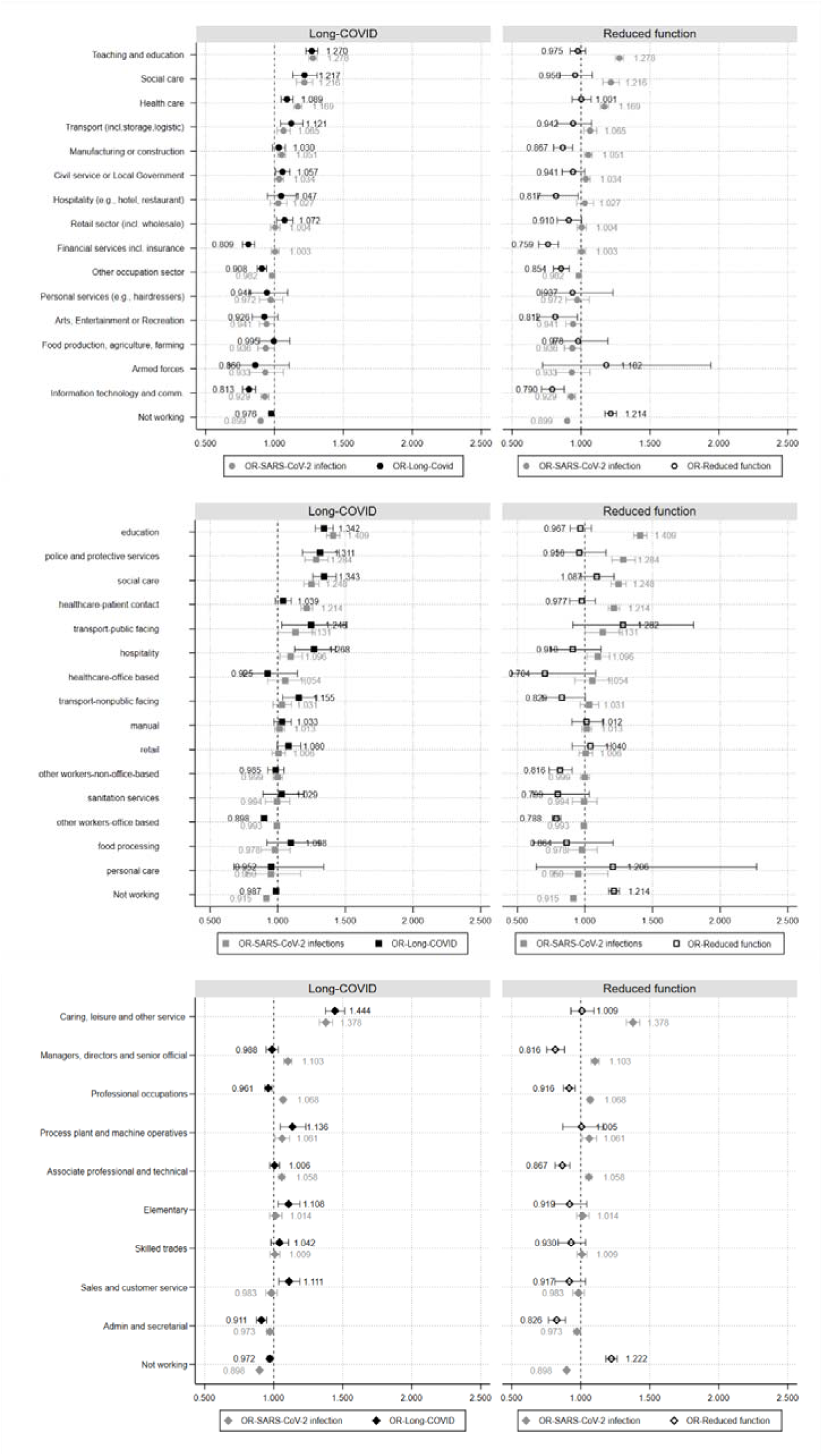
Odds (a-c) of having Long-COVID symptoms (left) and reduced function (right) by industry, occupational groupings and major SOC groups

By occupational group, in the unadjusted model most demonstrated higher likelihood of having long-COVID; however, for some CIs are wide (Table S7). Similar to analysis by industry, education (OR 1.53; CIs: 1.61-1.79) and Social Care (1.53; CIs: 1.45-1.60) occupations showed the highest risk. Manual, other workers-non-office based and office-based occupations, along with the non-working group showed ORs<1. After adjustment (Table S7, Figure 3b), education (1.34; CIs: 1.28-1.41) and social care (1.34; CIs: 1.26-1.43) remained the two groups with the highest relative risk and were closely followed by police and protective services (1.31; CIs 1.18-1.45), hospitality (127; CIs 1.13-1.43), and transport-public facing (1.25; CIs 1.03-1.51) or non-public facing (1.16; CIs 1.04-1.29). The group with the lowest ORs was office-based occupations (0.90; CIs: 0.88-0.92). All other groups with ORs≤1 have very wide confidence intervals. Prevalence of long-COVID by occupational group ranged from a low of 8.4% in other workers-office based, to a high of 12% in education (Table S9, Figure S1b).

Using major SOC group as the exposure, in the unadjusted model (Table S7), ORs for Caring, Leisure and other services stand out (OR 1.65, CIs: 1.58-1.73), compared to all other groups, followed by the sales and customers service, elementary, and process plant and machine operatives’ groups with ORs>1. In the remaining major SOC groups ORs were lower than one. After adjustment the category with the highest ORs remains the same; however, ORs were slightly attenuated. Minor changes were observed in all other groups (Table S7, Figure 3c). Admin and secretarial, professional occupations and those not working were less likely to report long-COVID symptoms. However, those in caring, leisure and other services (1.44, CIs: 1.38-1.53) still demonstrated substantially higher odds than average, followed by process plant and machine operatives (1.14, CIs: 1.05-1.23), sales and customer service (1.11, CIs: 1.04-1.19) and elementary occupations (1.11, CIs: 1.04-1.19). Finally, estimations for managers, directors and senior officials, and associate professional and technical occupations are uncertain. For most occupational groups prevalence of long-COVID was between 9 and 10%, with the exceptions of caring, leisure and other service occupations with 13% prevalence Analysis by major SOC group (Table S9, Figure S1c).

Comparing the likelihood of long-COVID with the likelihood of SARS-CoV-2 infection (Figures 3a-c) demonstrates similar patterns, i.e., industries with high risk of SARS-CoV-2 infection generally display high likelihood of long-COVID symptoms. In the majority of occupational groups similar patterns exist, with minor discrepancies for healthcare-office based where OR>1 for SARS-CoV-2 infection and OR<1 for long-COVID; and food processing and sanitation services where the opposite pattern is observed (Figure 3b). Using major SOC groups as the exposure displayed similar findings (Figure 3c), with the main exception of professional occupations, where likelihood of long-COVID is proportionally higher compared to SARS-CoV-2 infection. Some indication of different patterns between risk of infection and long-COVID were also observed in managers, directors and senior officials and sales occupations; however, in all cases confidence intervals are wide.

### Odds ratios (ORs) and prevalence (margins) of reduced function due to long-COVID

In order to examine the extent to which long-COVID symptoms impacted daily activities (i.e., proxy of severity) we used a reduced sample for all three exposure groups as analyses were restricted to those self-reporting at least one long-COVID symptom over the study period. Final analytical samples were 30 543 participants for industry, 25 888 participants for occupational groups and 28 979 participants for major SOC groups (Figure 1).

In the unadjusted model (Table S8) all industries, apart from healthcare (ORs 1.07; CIs: 1.00-1.14) and Social Care (1.07; CIs: 0.95-1.14), demonstrated lower odds of experiencing reduced function (i.e., reporting that daily activities were impacted a little or a lot) compared to the grand mean, while those not working had the highest odds (1.26; CIs: 1.23-2.03). Adjusting for all covariates attenuates ORs and in many cases confidence intervals are wide (e.g., Retail sector, food production and personal services); however, the trends remain stable (Table S8, Figure 3a).

Analysis by occupational groups (Table S8), similarly demonstrated that those not working have very high odds of reduced function (OR 1.25; CIs: 1.21-1.29); however, the highest odds are observed in the transport-public facing occupational group (1.29; CIs: 0.92-1.81), followed by personal (1.25; CIs: 0.67-2.32) and social care (1.23; CIs: 1.10-1.37) occupations. In all other groups ORs are less than one. Adjusting for all confounders generally attenuated the odds, but results retained the same patterns for magnitude and confidence intervals (Table S8, Figure 3a).

By major SOC group, in the unadjusted model (Table S8) for almost all groups it is less likely for someone to demonstrate reduced function due to long-COVID. The exceptions were the not working group (OR 1.27; CIs: 1.23-1.31) and the caring, leisure and other services (1.09; CIs: 1.01-1.18). Following adjustment, effect is attenuated for all groups (Table S8, Figure 3a).

Prevalence of reduced function by a ‘a little’ ranged between 47% and 49% for all three exposure groups (Table S9, Figure S2). Prevalence of reduced function by ‘a lot’, ranged from 17.1% to 21.7% by industry (for the arts, entertainment or recreation and teaching and education sectors, respectively), and 27.3%. for those not working. By occupational group, the lowest prevalence of reduced function by ‘a lot’ was observed for education (24.1%) and the highest for social care, personal care, and public-facing transport occupations. The latter were comparable to the prevalence for those not working (28.2%) (Table S9,Figure S2). By major SOC group, reduced function by ‘a lot’ ranged from 20.9% for managers, directors and senior officials and admin and secretarial to 24.4% for caring, leisure and other services and process plant and machine operatives.

### Sensitivity Panel Analysis - Relative risk (ORs) of long-COVID & reduced function due to long-COVID

Analysis run on the specified two additional samples, i.e. (i) confirmed PCR test or (ii) confirmed PCR test or prior report of infection (Figures S3-S4) did not show different patterns, confirming the robustness of our results.

For long-COVID odds ratios from the panel analysis were relatively larger compared to our main analysis for almost all industrial and occupational groups. The opposite was observed for major SOC groups, although the magnitude of the odds ratios were closer to those in our main sample. Similar patterns were observed for the outcome of reduced daily function (Tables S10-S12), suggesting assumptions about changes in occupation/occupational status over time are valid.

## Discussion

### Summary of findings

The prevalence of self-reported long-COVID increased through the study period. The social care and education industry (∼6%) and hospitality (∼4%) occupations recorded the highest prevalence. Industries and occupations with high levels of office-based working (e.g., IT), displayed lowest odds of long-COVID compared to more public facing industries, including teaching and education, social care, healthcare, civil service, retail and transport industries and occupations. By major SOC group, those in caring, leisure and other services (OR 1.44; CIs: 1.38-1.52) and education displayed (1.34; CIs: 1.28-1.41) substantially higher odds than average. The likelihood of reporting long-COVID symptoms followed the trend of risk of infection for most industries, occupational groups and SOC groups. The exception was for professional occupations, where relative risk of reporting long-COVID symptoms was higher than that of infections. Indication of differing patterns between infection and long-COVID were also observed in other occupational groups, but results uncertainty was high. Long-COVID symptoms impacted almost half of all affected participants’ ability to do daily activities by ‘a little’. Participants in healthcare and social care industries were most impacted by ‘a lot’. Similarly, occupational groups most affected were public-facing occupations with those working in transport occupations having the highest prevalence of all (30%), followed by the not working (28.1%) and personal care (27.4%). For SOC groups the highest prevalence (24%) was observed in caring, leisure and other services and process plant and machine operatives.

### Findings in context with previous literature

To our knowledge, no other studies examine long-COVID across industries and/or occupational groups for direct comparisons. However, previous work estimated that ∼10% of people infected with Covid-19 may have significant post-acute or chronic symptoms persisting >12 weeks (23) and that a substantial proportion of these will have symptoms that are disabling and involve prolonged absence from work (24). These findings are in line with our results (23 24). A study in adult patients with COVID-19 attending a hospital showed that post-acute COVID-19 syndrome was detected in a half of COVID-19 survivors (25). These are higher than our results but were an older cohort and probably represent a group with severe COVID-19 infection (25). Vulnerable groups are disproportionately represented among essential workers (e.g., bus drivers, allied health professionals) (26 27). In addition to being at a greater risk of infection (3 4 28), they may also face a disproportionate burden of long-COVID. Our findings suggest that workers in social care, education, hospitality, food production and transport have increased odds of long-COVID. Moreover, groups likely to have high levels of long-COVID include those with a high risk of SARS-CoV2 infection (26), with the exception of professional occupations.

### Strengths & Limitations

Our study has several strengths. We used a large nationally representative dataset to examine prevalence and risk of long-COVID across industries, and occupations, which fills an important gap in the literature. Our analysis included a relatively long study period (Feb2021-Apr2022), allowing us to examine trends of over time. Some limitations should be noted. Any analysis by occupation/sector requires aggregation of different types of workers in groups. It is possible that analyses mask differences within particular sectors or occupations. In our main analyses we utilised occupational status at the beginning of the study period, which may mask effects due to changes in employment status. However, our sensitivity analysis using a panel dataset addressed this. At the time of data collection, there was no universal definition of long-COVID, and the results reported rely on self-reported measures which are prone to bias. It is possible that bias in this measure is related to occupation – for example healthcare workers increased health knowledge may mean that they self-diagnose long-COVID differently. The CIS recording of COVID-19 infections does not include infections occurring between visits and missed visits may be related to occupation e.g., shift-workers being unavailable at the CIS visit. It is not possible to assess the reason(s) participant are not working. Therefore, we cannot examine or exclude the possibility that some participants are not working due to long-COVID symptoms.

### Conclusions

The SARS-CoV-2 pandemic created new challenges for workers, employers, occupational and public health. Long-COVID is compounding to these challenges, mainly related to sustaining employment and return-to-work for those affected. Our findings show that long-COVID differences are evident across industries and occupations. Professional occupations, including jobs in teaching and education, IT, welfare and healthcare showed that the risk of infection may not be the only driving force. Our findings of increased risk of long-COVID in the health and social care industries support the findings that some health conditions are prescribed as industrial diseases (7) and highlight other sectors and occupations that require further attention. Scientific evidence is essential to understand the mechanisms resulting in long-COVID, how occupation influences prevalence and severity and whether working conditions affect the risk of developing long-COVID or interact with long-COVID to increase the impact on activities.

## Data Availability

ONS CIS data can be accessed only by researchers who are Office of National Statistics (ONS) accredited researchers. Researchers can apply for accreditation through the Research Accreditation Service. Access is through the Secure Research Service (SRS) and approved on a project basis. For further details see: https://www.ons.gov.uk/aboutus/whatwedo/statistics/requestingstatistics/approvedresearcherscheme.

## Contributors

SR is the principal investigator. All authors contributed to the design of the proposal and study. TK lead and conducted the statistical analysis and was supported by ED and SR. TK and ED drafted the manuscript. All authors contributed to the interpretation of the results, critically revised the paper and agreed on the final version for submission.

## Disclaimer

This work contains statistical data from ONS which is Crown Copyright. The use of the ONS statistical data in this work does not imply the endorsement of the ONS in relation to the interpretation or analysis of the statistical data. This work uses research datasets which may not exactly reproduce National Statistics aggregates.

## Ethics approval

The COVID-19 Infection Survey received ethical approval from the South-Central Berkshire B Research Ethics Committee (20/SC/0195). All participants provided informed consent. For use of this data for this project statistics authority self-assessment classified the study as low risk. This assessment was approved by the Office for National Statistics (ONS) Research Accreditation Panel.

## Patient and public involvement

Participants were not involved in the design and implementation of the study or in setting research questions and the outcome measures. No participants were asked to advise on interpretation or writing up of results.

