## Supplementary Material for "Occupational differences in the prevalence and severity of long-COVID: Analysis of the ONS Coronavirus (COVID-19) Infection Survey"

**Employment status coding**

The working group included those employed and self-employed XYZ and the not working those who reported being students, furloughed (temporarily not working), unemployed, retired, long-term sick etc, those that are employed or self-employed but currently are not working for reason other than furlough, those looking for paid work being able start and those not working but also not looking for work. First variable used: work_status_clean (0:Not working, 1: Working, 2: Student (**0 and 2 were moved to Not working)**

Second variable used: work_status (1: Employed, 2: Self-employed, 3:Furloughed (temporarily not working), 4: Not working (unemployed, retired, long-term sick etc), 5: Student (**3,4 and 5 were moved to Not working)**

Third variable used: work_status_v1: (1: Employed and currently working, 2: Employed and currently not working, 3: Self-Employed and currently working, 4: Self-Employed and currently not working, 5: Looking for paid work and able to start, 6: Not working and not looking for work, 7: Retired, 8: Child under 5y not attending child care, 9: Child under 5y attending child care, 10: 5y old and older in full-time education (**2,4 and** **5 to 10 were moved to Not working)**

Fourth variable used: work_status_v2: (1: Employed and currently working, 2: Employed and currently not working, 3: Self-Employed and currently working, 4: Self-Employed and currently not working, 5: Looking for paid work and able to start, 6: Not working and not looking for work, 7: Retired, 8: Child under 4-5y not attending child care, 9: Child under 4-5y attending child care, 10: 5y old and older at school/home-school, 11: Attending college or FE (including if temporary absent), 12: Attending university (including if temporary absent (**2,4 and 5 to 12 were moved to Not working)**

**Handling of duplicates**

We identified multiple duplicate observations (947) for individuals on the same date (e.g., for additional blood tests). To address this, first, we replaced Follow up (1) to First visit (0) where visit number equalled zero and visit date was the same (105 observations). Then, seven observations were changed to follow up visit value (1) from First visit (0) as these occurred after the original First visit. Finally, we deleted the remaining duplicates (477 observations (), creating our final dataset with one observation per individual. This analysis allowed for variables’ values to change across time.

Occupations were grouped based on a consensus building exercise between members of the research team and other experts that expanded on the essential workers grouping previously used in the Mutambudzi et al study (1) to reflect potential exposures during the initial phase of the pandemic. Using SOC 2010 4 digit codes, the consensus produced eleven main groups (Occ_groups; Table S1) which could be further split to 15 subgroups (Occ_subgroups; Table S 1). The new categories were mapped onto the COVID-19-JEM (Table S 1).

Table S 1 Occupational groupings and mapping with Occupational Job Exposure Matrix (JEM) domains*†.

| SOC2010 code | SOC2010 name | Occ_ groups | Occ_subgroups | JEM Domain 1 | JEM Domain 2 | JEM Domain 3 | JEM Domain 4 | JEM Domain 5 | JEM Domain 6 | JEM Domain 7 | JEM Domain 8 |
| --- | --- | --- | --- | --- | --- | --- | --- | --- | --- | --- | --- |
| 1115 | Chief executives and senior officials | other workers | other workers-office based | 0 | 0 | 0 | 0 | 0 | 0 | 1 | 0 |
| 1116 | Elected officers and representatives | other workers | other workers-office based | 2 | 1 | 1 | 3 | 1 | 2 | 1 | 0 |
| 1121 | Production managers and directors in manufacturing | other workers | other workers-office based | 3 | 1 | 1 | 3 | 1 | 2 | 1 | 1 |
| 1122 | Production managers and directors in construction | other workers | other workers-office based | 3 | 1 | 1 | 2 | 1 | 2 | 1 | 0 |
| 1123 | Production managers and directors in mining and energy | other workers | other workers-office based | 3 | 1 | 1 | 3 | 1 | 2 | 1 | 0 |
| 1131 | Financial managers and directors | other workers | other workers-office based | 0 | 0 | 0 | 0 | 0 | 0 | 1 | 0 |
| 1132 | Marketing and sales directors | other workers | other workers-office based | 0 | 0 | 0 | 0 | 0 | 0 | 1 | 0 |
| 1133 | Purchasing managers and directors | other workers | other workers-office based | 2 | 1 | 1 | 3 | 1 | 2 | 1 | 0 |
| 1134 | Advertising and public relations directors | other workers | other workers-office based | 0 | 0 | 0 | 0 | 0 | 0 | 1 | 0 |
| 1135 | Human resource managers and directors | other workers | other workers-office based | 0 | 0 | 0 | 0 | 0 | 0 | 1 | 0 |
| 1136 | Information technology and telecommunications directors | other workers | other workers-office based | 0 | 0 | 0 | 0 | 0 | 0 | 1 | 0 |
| 1139 | Functional managers and directors n.e.c. | other workers | other workers-office based | 0 | 0 | 0 | 0 | 0 | 0 | 1 | 0 |
| 1150 | Financial institution managers and directors | other workers | other workers-office based | 2 | 2 | 2 | 3 | 1 | 2 | 2 | 1 |
| 1161 | Managers and directors in transport and distribution | other workers | other workers-office based | 2 | 1 | 1 | 3 | 1 | 2 | 1 | 1 |
| 1162 | Managers and directors in storage and warehousing | other workers | other workers-office based | 2 | 1 | 1 | 3 | 1 | 2 | 1 | 1 |
| 1171 | Officers in armed forces | police and protective services | police and protective services | 3 | 1 | 1 | 3 | 1 | 2 | 0 | 0 |
| 1172 | Senior police officers | police and protective services | police and protective services | 2 | 2 | 2 | 2 | 2 | 2 | 0 | 0 |
| 1173 | Senior officers in fire, ambulance, prison and related services | police and protective services | police and protective services | 3 | 0 | 1 | 3 | 1 | 2 | 0 | 0 |
| 1181 | Health services and public health managers and directors | healthcare | healthcare-office based | 3 | 3 | 3 | 3 | 1 | 2 | 1 | 1 |
| 1184 | Social services managers and directors | social and education | social care | 1 | 0 | 1 | 3 | 1 | 2 | 1 | 0 |
| 1190 | Managers and directors in retail and wholesale | retail | retail | 3 | 3 | 2 | 3 | 1 | 2 | 1 | 0 |
| 1211 | Managers and proprietors in agriculture and horticulture | other workers | other workers-non-office based | 1 | 0 | 1 | 2 | 1 | 2 | 1 | 0 |
| 1213 | Managers and proprietors in forestry, fishing and related services | other workers | other workers-non-office based | 1 | 0 | 1 | 2 | 1 | 2 | 1 | 0 |
| 1221 | Hotel and accommodation managers and proprietors | hospitality | hospitality | 3 | 3 | 2 | 3 | 1 | 2 | 2 | 0 |
| 1223 | Restaurant and catering establishment managers and proprietors | hospitality | hospitality | 2 | 3 | 2 | 3 | 1 | 2 | 2 | 1 |
| 1224 | Publicans and managers of licensed premises | hospitality | hospitality | 3 | 3 | 2 | 3 | 1 | 2 | 2 | 1 |
| 1225 | Leisure and sports managers | other workers | other workers-office based | 2 | 3 | 2 | 2 | 1 | 2 | 2 | 1 |
| 1226 | Travel agency managers and proprietors | other workers | other workers-office based | 2 | 3 | 2 | 3 | 1 | 2 | 2 | 1 |
| 1241 | Health care practice managers | healthcare | healthcare-office based | 3 | 2 | 3 | 3 | 1 | 1 | 1 | 1 |
| 1242 | Residential, day and domiciliary care managers and proprietors | social and education | social care | 3 | 3 | 3 | 3 | 1 | 2 | 1 | 0 |
| 1251 | Property, housing and estate managers | other workers | other workers-non-office based | 2 | 3 | 2 | 3 | 1 | 2 | 1 | 0 |
| 1252 | Garage managers and proprietors | other workers | other workers-non-office based | 2 | 3 | 2 | 3 | 1 | 2 | 1 | 0 |
| 1253 | Hairdressing and beauty salon managers and proprietors | personal care | personal care | 2 | 3 | 2 | 3 | 3 | 1 | 1 | 0 |
| 1254 | Shopkeepers and proprietors – wholesale and retail | retail | retail | 3 | 3 | 2 | 3 | 2 | 1 | 1 | 0 |
| 1255 | Waste disposal and environmental services managers | sanitation services | sanitation services | 3 | 1 | 1 | 3 | 1 | 2 | 1 | 0 |
| 1259 | Managers and proprietors in other services n.e.c. | other workers | other workers-office based | 2 | 3 | 2 | 3 | 1 | 2 | 1 | 0 |
| 2111 | Chemical scientists | other workers | other workers-non-office based | 1 | 1 | 1 | 3 | 1 | 2 | 2 | 0 |
| 2112 | Biological scientists and biochemists | other workers | other workers-non-office based | 1 | 1 | 1 | 2 | 1 | 2 | 2 | 1 |
| 2113 | Physical scientists | other workers | other workers-non-office based | 0 | 0 | 0 | 0 | 0 | 0 | 2 | 0 |
| 2114 | Social and humanities scientists | other workers | other workers-non-office based | 0 | 0 | 0 | 0 | 0 | 0 | 2 | 1 |
| 2119 | Natural and social science professionals n.e.c. | other workers | other workers-office based | 0 | 0 | 0 | 0 | 0 | 0 | 2 | 1 |
| 2121 | Civil engineers | other workers | other workers-non-office based | 1 | 1 | 1 | 3 | 1 | 2 | 1 | 0 |
| 2122 | Mechanical engineers | other workers | other workers-non-office based | 2 | 1 | 1 | 3 | 1 | 2 | 1 | 1 |
| 2123 | Electrical engineers | other workers | other workers-non-office based | 1 | 1 | 1 | 3 | 1 | 2 | 1 | 0 |
| 2124 | Electronics engineers | other workers | other workers-non-office based | 1 | 1 | 1 | 3 | 1 | 2 | 1 | 0 |
| 2126 | Design and development engineers | other workers | other workers-non-office based | 1 | 1 | 1 | 3 | 1 | 2 | 1 | 0 |
| 2127 | Production and process engineers | other workers | other workers-non-office based | 2 | 1 | 1 | 3 | 1 | 2 | 1 | 1 |
| 2129 | Engineering professionals n.e.c. | other workers | other workers-non-office based | 1 | 1 | 1 | 3 | 1 | 2 | 1 | 0 |
| 2133 | IT specialist managers | other workers | other workers-office based | 0 | 0 | 0 | 0 | 0 | 0 | 2 | 0 |
| 2134 | IT project and programme managers | other workers | other workers-office based | 0 | 0 | 0 | 0 | 0 | 0 | 2 | 1 |
| 2135 | IT business analysts, architects and systems designers | other workers | other workers-office based | 0 | 0 | 0 | 0 | 0 | 0 | 2 | 0 |
| 2136 | Programmers and software development professionals | other workers | other workers-office based | 0 | 0 | 0 | 0 | 0 | 0 | 2 | 0 |
| 2137 | Web design and development professionals | other workers | other workers-office based | 0 | 0 | 0 | 0 | 0 | 0 | 2 | 0 |
| 2139 | Information technology and telecommunications professionals n.e.c. | other workers | other workers-office based | 0 | 0 | 0 | 0 | 0 | 0 | 2 | 0 |
| 2141 | Conservation professionals | other workers | other workers-non-office based | 1 | 1 | 1 | 2 | 1 | 2 | 2 | 0 |
| 2142 | Environment professionals | other workers | other workers-non-office based | 1 | 1 | 1 | 2 | 1 | 2 | 2 | 0 |
| 2150 | Air traffic controllers | other workers | other workers-office based | 0 | 0 | 0 | 0 | 0 | 0 | 2 | 1 |
| 2211 | Medical practitioners | healthcare | healthcare-patient contact | 2 | 3 | 3 | 3 | 2 | 1 | 2 | 1 |
| 2212 | Psychologists | healthcare | healthcare-patient contact | 2 | 2 | 2 | 3 | 1 | 1 | 2 | 0 |
| 2213 | Pharmacists | healthcare | healthcare-patient contact | 3 | 2 | 3 | 3 | 1 | 1 | 2 | 1 |
| 2214 | Ophthalmic opticians | healthcare | healthcare-patient contact | 2 | 3 | 2 | 3 | 3 | 1 | 2 | 0 |
| 2215 | Dental practitioners | healthcare | healthcare-patient contact | 2 | 3 | 2 | 3 | 3 | 1 | 2 | 0 |
| 2216 | Veterinarians | other workers | other workers-non-office based | 2 | 3 | 2 | 3 | 2 | 1 | 2 | 0 |
| 2217 | Medical radiographers | healthcare | healthcare-patient contact | 2 | 3 | 3 | 3 | 3 | 1 | 2 | 1 |
| 2218 | Podiatrists | healthcare | healthcare-patient contact | 3 | 3 | 3 | 3 | 2 | 1 | 2 | 0 |
| 2219 | Health professionals n.e.c. | healthcare | healthcare-patient contact | 3 | 3 | 3 | 3 | 2 | 1 | 2 | 0 |
| 2221 | Physiotherapists | healthcare | healthcare-patient contact | 2 | 3 | 2 | 3 | 3 | 1 | 1 | 1 |
| 2222 | Occupational therapists | healthcare | healthcare-patient contact | 3 | 3 | 3 | 3 | 2 | 1 | 1 | 0 |
| 2223 | Speech and language therapists | healthcare | healthcare-patient contact | 2 | 2 | 2 | 3 | 2 | 2 | 1 | 0 |
| 2229 | Therapy professionals n.e.c. | healthcare | healthcare-patient contact | 2 | 2 | 2 | 3 | 1 | 1 | 1 | 1 |
| 2231 | Nurses | healthcare | healthcare-patient contact | 3 | 3 | 3 | 3 | 3 | 1 | 1 | 1 |
| 2232 | Midwives | healthcare | healthcare-patient contact | 2 | 3 | 2 | 3 | 3 | 1 | 1 | 1 |
| 2311 | Higher education teaching professionals | social and education | education | 0 | 0 | 0 | 0 | 0 | 0 | 2 | 1 |
| 2312 | Further education teaching professionals | social and education | education | 3 | 3 | 2 | 3 | 2 | 2 | 2 | 1 |
| 2314 | Secondary education teaching professionals | social and education | education | 3 | 3 | 2 | 3 | 2 | 2 | 2 | 1 |
| 2315 | Primary and nursery education teaching professionals | social and education | education | 2 | 2 | 2 | 3 | 2 | 2 | 2 | 0 |
| 2316 | Special needs education teaching professionals | social and education | education | 2 | 2 | 2 | 3 | 2 | 2 | 2 | 1 |
| 2317 | Senior professionals of educational establishments | social and education | education | 3 | 1 | 1 | 3 | 1 | 2 | 2 | 0 |
| 2318 | Education advisers and school inspectors | social and education | education | 1 | 1 | 2 | 3 | 1 | 2 | 2 | 0 |
| 2319 | Teaching and other educational professionals n.e.c. | social and education | education | 2 | 2 | 2 | 3 | 1 | 2 | 2 | 1 |
| 2412 | Barristers and judges | other workers | other workers-office based | 2 | 3 | 2 | 3 | 1 | 2 | 1 | 1 |
| 2413 | Solicitors | other workers | other workers-office based | 2 | 3 | 2 | 3 | 1 | 2 | 1 | 0 |
| 2419 | Legal professionals n.e.c. | other workers | other workers-office based | 2 | 3 | 2 | 3 | 1 | 2 | 1 | 0 |
| 2421 | Chartered and certified accountants | other workers | other workers-office based | 1 | 2 | 2 | 3 | 1 | 2 | 2 | 0 |
| 2423 | Management consultants and business analysts | other workers | other workers-office based | 0 | 0 | 0 | 0 | 0 | 0 | 2 | 1 |
| 2424 | Business and financial project management professionals | other workers | other workers-office based | 0 | 0 | 0 | 0 | 0 | 0 | 2 | 0 |
| 2425 | Actuaries, economists and statisticians | other workers | other workers-office based | 0 | 0 | 0 | 0 | 0 | 0 | 2 | 0 |
| 2426 | Business and related research professionals | other workers | other workers-office based | 0 | 0 | 0 | 0 | 0 | 0 | 2 | 0 |
| 2429 | Business, research and administrative professionals n.e.c. | other workers | other workers-office based | 1 | 0 | 1 | 3 | 1 | 2 | 2 | 1 |
| 2431 | Architects | other workers | other workers-office based | 1 | 0 | 1 | 2 | 1 | 2 | 1 | 1 |
| 2432 | Town planning officers | other workers | other workers-office based | 0 | 0 | 0 | 0 | 0 | 0 | 1 | 0 |
| 2433 | Quantity surveyors | other workers | other workers-office based | 1 | 1 | 1 | 3 | 1 | 2 | 1 | 1 |
| 2434 | Chartered surveyors | other workers | other workers-office based | 0 | 0 | 0 | 0 | 0 | 0 | 1 | 0 |
| 2435 | Chartered architectural technologists | other workers | other workers-office based | 1 | 0 | 1 | 2 | 1 | 2 | 1 | 0 |
| 2436 | Construction project managers and related professionals | other workers | other workers-non-office based | 3 | 1 | 1 | 2 | 1 | 2 | 1 | 1 |
| 2442 | Social workers | social and education | social care | 3 | 3 | 2 | 3 | 2 | 1 | 1 | 0 |
| 2443 | Probation officers | social and education | social care | 3 | 3 | 2 | 3 | 2 | 1 | 1 | 0 |
| 2444 | Clergy | social and education | social care | 3 | 3 | 2 | 3 | 2 | 2 | 1 | 0 |
| 2449 | Welfare professionals n.e.c. | social and education | social care | 3 | 3 | 2 | 3 | 2 | 1 | 1 | 0 |
| 2451 | Librarians | other workers | other workers-office based | 2 | 3 | 2 | 3 | 1 | 2 | 1 | 1 |
| 2452 | Archivists and curators | other workers | other workers-office based | 1 | 1 | 1 | 3 | 1 | 2 | 1 | 1 |
| 2461 | Quality control and planning engineers | other workers | other workers-non-office based | 1 | 1 | 1 | 3 | 1 | 2 | 1 | 0 |
| 2462 | Quality assurance and regulatory professionals | other workers | other workers-office based | 0 | 0 | 0 | 0 | 0 | 0 | 1 | 0 |
| 2463 | Environmental health professionals | other workers | other workers-non-office based | 2 | 2 | 2 | 3 | 1 | 2 | 1 | 0 |
| 2471 | Journalists, newspaper and periodical editors | other workers | other workers-office based | 1 | 0 | 2 | 3 | 1 | 2 | 2 | 1 |
| 2472 | Public relations professionals | other workers | other workers-office based | 0 | 0 | 0 | 0 | 0 | 0 | 2 | 1 |
| 2473 | Advertising accounts managers and creative directors | other workers | other workers-office based | 0 | 0 | 0 | 0 | 0 | 0 | 2 | 0 |
| 3111 | Laboratory technicians | other workers | other workers-non-office based | 2 | 1 | 1 | 3 | 1 | 2 | 1 | 1 |
| 3112 | Electrical and electronics technicians | other workers | other workers-non-office based | 2 | 1 | 1 | 3 | 1 | 2 | 1 | 0 |
| 3113 | Engineering technicians | other workers | other workers-non-office based | 2 | 1 | 1 | 3 | 1 | 2 | 1 | 0 |
| 3114 | Building and civil engineering technicians | other workers | other workers-non-office based | 2 | 1 | 1 | 3 | 1 | 2 | 1 | 0 |
| 3115 | Quality assurance technicians | other workers | other workers-non-office based | 2 | 1 | 1 | 3 | 1 | 2 | 1 | 1 |
| 3116 | Planning, process and production technicians | other workers | other workers-non-office based | 2 | 1 | 1 | 3 | 1 | 2 | 1 | 0 |
| 3119 | Science, engineering and production technicians n.e.c. | other workers | other workers-non-office based | 2 | 1 | 1 | 3 | 1 | 2 | 1 | 0 |
| 3121 | Architectural and town planning technicians | other workers | other workers-non-office based | 2 | 1 | 1 | 3 | 1 | 2 | 2 | 0 |
| 3122 | Draughtspersons | other workers | other workers-non-office based | 0 | 0 | 0 | 0 | 0 | 0 | 2 | 0 |
| 3131 | IT operations technicians | other workers | other workers-office based | 0 | 0 | 0 | 0 | 0 | 0 | 1 | 0 |
| 3132 | IT user support technicians | other workers | other workers-office based | 1 | 0 | 1 | 3 | 1 | 2 | 1 | 0 |
| 3213 | Paramedics | healthcare | healthcare-patient contact | 2 | 3 | 3 | 2 | 3 | 1 | 1 | 0 |
| 3216 | Dispensing opticians | healthcare | healthcare-patient contact | 2 | 3 | 2 | 3 | 2 | 1 | 1 | 1 |
| 3217 | Pharmaceutical technicians | healthcare | healthcare-patient contact | 3 | 3 | 3 | 3 | 1 | 1 | 1 | 0 |
| 3218 | Medical and dental technicians | healthcare | healthcare-patient contact | 2 | 3 | 3 | 3 | 3 | 1 | 1 | 1 |
| 3219 | Health associate professionals n.e.c. | healthcare | healthcare-patient contact | 2 | 3 | 2 | 3 | 2 | 1 | 1 | 0 |
| 3231 | Youth and community workers | social and education | social care | 2 | 2 | 2 | 3 | 2 | 2 | 1 | 1 |
| 3233 | Child and early years officers | social and education | social care | 2 | 2 | 2 | 3 | 2 | 2 | 1 | 1 |
| 3234 | Housing officers | social and education | social care | 2 | 2 | 2 | 3 | 2 | 2 | 1 | 1 |
| 3235 | Counsellors | social and education | social care | 3 | 3 | 2 | 3 | 2 | 1 | 1 | 0 |
| 3239 | Welfare and housing associate professionals n.e.c. | social and education | social care | 2 | 2 | 2 | 3 | 2 | 2 | 1 | 1 |
| 3311 | NCOs and other ranks | police and protective services | police and protective services | 3 | 1 | 1 | 3 | 1 | 2 | 1 | 0 |
| 3312 | Police officers (sergeant and below) | police and protective services | police and protective services | 3 | 1 | 2 | 2 | 3 | 2 | 1 | 0 |
| 3313 | Fire service officers (watch manager and below) | police and protective services | police and protective services | 2 | 1 | 2 | 2 | 2 | 2 | 1 | 0 |
| 3314 | Prison service officers (below principal officer) | police and protective services | police and protective services | 3 | 3 | 2 | 3 | 3 | 2 | 1 | 0 |
| 3315 | Police community support officers | police and protective services | police and protective services | 3 | 1 | 2 | 2 | 3 | 2 | 1 | 0 |
| 3319 | Protective service associate professionals n.e.c. | police and protective services | police and protective services | 3 | 3 | 2 | 3 | 1 | 2 | 1 | 1 |
| 3411 | Artists | other workers | other workers-office based | 0 | 0 | 0 | 0 | 0 | 0 | 1 | 0 |
| 3412 | Authors, writers and translators | other workers | other workers-office based | 0 | 0 | 0 | 0 | 0 | 0 | 1 | 1 |
| 3413 | Actors, entertainers and presenters | other workers | other workers-non-office based | 2 | 1 | 1 | 3 | 3 | 3 | 1 | 1 |
| 3414 | Dancers and choreographers | other workers | other workers-non-office based | 2 | 2 | 2 | 3 | 1 | 2 | 1 | 1 |
| 3415 | Musicians | other workers | other workers-non-office based | 1 | 1 | 1 | 3 | 1 | 3 | 1 | 1 |
| 3416 | Arts officers, producers and directors | other workers | other workers-non-office based | 2 | 1 | 1 | 3 | 2 | 3 | 1 | 0 |
| 3417 | Photographers, audio-visual and broadcasting equipment operators | other workers | other workers-non-office based | 1 | 0 | 2 | 2 | 1 | 2 | 1 | 1 |
| 3421 | Graphic designers | other workers | other workers-office based | 0 | 0 | 0 | 0 | 0 | 0 | 2 | 1 |
| 3422 | Product, clothing and related designers | other workers | other workers-office based | 1 | 1 | 1 | 3 | 1 | 2 | 2 | 1 |
| 3441 | Sports players | other workers | other workers-non-office based | 2 | 1 | 1 | 2 | 3 | 3 | 1 | 1 |
| 3442 | Sports coaches, instructors and officials | other workers | other workers-non-office based | 2 | 1 | 1 | 2 | 2 | 3 | 1 | 1 |
| 3443 | Fitness instructors | other workers | other workers-non-office based | 2 | 3 | 2 | 2 | 2 | 2 | 1 | 1 |
| 3511 | Air traffic controllers | other workers | other workers-office based | 2 | 1 | 1 | 3 | 1 | 2 | 1 | 0 |
| 3512 | Aircraft pilots and flight engineers | transport | transport-public facing | 3 | 1 | 2 | 3 | 3 | 2 | 1 | 0 |
| 3513 | Ship and hovercraft officers | transport | transport-nonpublic facing | 1 | 1 | 1 | 3 | 3 | 2 | 1 | 0 |
| 3520 | Legal associate professionals | other workers | other workers-office based | 2 | 3 | 2 | 2 | 1 | 2 | 1 | 1 |
| 3531 | Estimators, valuers and assessors | other workers | other workers-office based | 1 | 2 | 2 | 3 | 1 | 2 | 1 | 1 |
| 3532 | Brokers | other workers | other workers-office based | 2 | 0 | 1 | 3 | 1 | 2 | 1 | 0 |
| 3533 | Insurance underwriters | other workers | other workers-office based | 0 | 0 | 0 | 0 | 0 | 0 | 1 | 0 |
| 3534 | Finance and investment analysts and advisers | other workers | other workers-office based | 0 | 0 | 0 | 0 | 0 | 0 | 1 | 1 |
| 3535 | Taxation experts | other workers | other workers-office based | 1 | 2 | 2 | 3 | 1 | 2 | 1 | 0 |
| 3536 | Importers and exporters | other workers | other workers-office based | 2 | 0 | 1 | 3 | 1 | 2 | 1 | 0 |
| 3537 | Financial and accounting technicians | other workers | other workers-office based | 1 | 2 | 2 | 3 | 1 | 2 | 1 | 0 |
| 3538 | Financial accounts managers | other workers | other workers-office based | 0 | 0 | 0 | 0 | 0 | 0 | 1 | 0 |
| 3539 | Business and related associate professionals n.e.c. | other workers | other workers-office based | 0 | 0 | 0 | 0 | 0 | 0 | 1 | 0 |
| 3541 | Buyers and procurement officers | other workers | other workers-office based | 0 | 0 | 0 | 0 | 0 | 0 | 2 | 0 |
| 3542 | Business sales executives | other workers | other workers-office based | 0 | 0 | 0 | 0 | 0 | 0 | 2 | 0 |
| 3543 | Marketing associate professionals | other workers | other workers-office based | 0 | 0 | 0 | 0 | 0 | 0 | 2 | 0 |
| 3544 | Estate agents and auctioneers | other workers | other workers-office based | 2 | 2 | 2 | 3 | 1 | 2 | 2 | 1 |
| 3545 | Sales accounts and business development managers | other workers | other workers-office based | 1 | 2 | 2 | 3 | 1 | 2 | 2 | 0 |
| 3546 | Conference and exhibition managers and organisers | other workers | other workers-office based | 0 | 0 | 0 | 0 | 0 | 0 | 2 | 1 |
| 3550 | Conservation and environmental associate professionals | other workers | other workers-non-office based | 1 | 1 | 1 | 1 | 1 | 2 | 1 | 0 |
| 3561 | Public services associate professionals | other workers | other workers-office based | 1 | 0 | 1 | 3 | 1 | 2 | 1 | 1 |
| 3562 | Human resources and industrial relations officers | other workers | other workers-office based | 0 | 0 | 0 | 0 | 0 | 0 | 1 | 0 |
| 3563 | Vocational and industrial trainers and instructors | other workers | other workers-office based | 0 | 0 | 0 | 0 | 0 | 0 | 1 | 0 |
| 3564 | Careers advisers and vocational guidance specialists | other workers | other workers-office based | 0 | 0 | 0 | 0 | 0 | 0 | 1 | 0 |
| 3565 | Inspectors of standards and regulations | other workers | other workers-non-office based | 1 | 0 | 1 | 3 | 1 | 2 | 1 | 0 |
| 3567 | Health and safety officers | other workers | other workers-non-office based | 2 | 2 | 2 | 3 | 1 | 2 | 1 | 1 |
| 4112 | National government administrative occupations | other workers | other workers-office based | 0 | 0 | 0 | 0 | 0 | 0 | 1 | 0 |
| 4113 | Local government administrative occupations | other workers | other workers-office based | 2 | 1 | 1 | 3 | 1 | 2 | 1 | 1 |
| 4114 | Officers of non-governmental organisations | other workers | other workers-office based | 0 | 0 | 0 | 0 | 0 | 0 | 1 | 0 |
| 4121 | Credit controllers | other workers | other workers-office based | 0 | 0 | 0 | 0 | 0 | 0 | 1 | 0 |
| 4122 | Book-keepers, payroll managers and wages clerks | other workers | other workers-office based | 0 | 0 | 0 | 0 | 0 | 0 | 1 | 0 |
| 4123 | Bank and post office clerks | other workers | other workers-office based | 3 | 3 | 2 | 3 | 1 | 1 | 1 | 1 |
| 4124 | Finance officers | other workers | other workers-office based | 0 | 0 | 0 | 0 | 0 | 0 | 1 | 0 |
| 4129 | Financial administrative occupations n.e.c. | other workers | other workers-office based | 3 | 3 | 2 | 3 | 1 | 1 | 1 | 1 |
| 4131 | Records clerks and assistants | other workers | other workers-office based | 0 | 0 | 0 | 0 | 0 | 0 | 1 | 0 |
| 4132 | Pensions and insurance clerks and assistants | other workers | other workers-office based | 0 | 0 | 0 | 0 | 0 | 0 | 1 | 0 |
| 4133 | Stock control clerks and assistants | retail | retail | 1 | 1 | 1 | 3 | 1 | 2 | 1 | 1 |
| 4134 | Transport and distribution clerks and assistants | transport | transport-nonpublic facing | 2 | 0 | 1 | 3 | 1 | 2 | 1 | 1 |
| 4135 | Library clerks and assistants | other workers | other workers-office based | 3 | 3 | 2 | 3 | 1 | 2 | 1 | 0 |
| 4138 | Human resources administrative occupations | other workers | other workers-office based | 0 | 0 | 0 | 0 | 0 | 0 | 1 | 0 |
| 4151 | Sales administrators | retail | retail | 0 | 0 | 0 | 0 | 0 | 0 | 1 | 1 |
| 4159 | Other administrative occupations n.e.c. | other workers | other workers-office based | 0 | 0 | 0 | 0 | 0 | 0 | 1 | 1 |
| 4161 | Office managers | other workers | other workers-office based | 2 | 0 | 1 | 3 | 1 | 2 | 1 | 0 |
| 4162 | Office supervisors | other workers | other workers-office based | 2 | 0 | 1 | 3 | 1 | 2 | 1 | 1 |
| 4211 | Medical secretaries | healthcare | healthcare-office based | 3 | 2 | 3 | 3 | 1 | 1 | 1 | 1 |
| 4212 | Legal secretaries | other workers | other workers-office based | 1 | 1 | 1 | 3 | 1 | 2 | 1 | 0 |
| 4213 | School secretaries | social and education | education | 2 | 1 | 1 | 3 | 2 | 2 | 1 | 0 |
| 4214 | Company secretaries | other workers | other workers-office based | 0 | 0 | 0 | 0 | 0 | 0 | 1 | 0 |
| 4215 | Personal assistants and other secretaries | other workers | other workers-office based | 0 | 0 | 0 | 0 | 0 | 0 | 1 | 0 |
| 4216 | Receptionists | other workers | other workers-office based | 2 | 3 | 2 | 3 | 1 | 1 | 1 | 1 |
| 4217 | Typists and related keyboard occupations | other workers | other workers-office based | 0 | 0 | 0 | 0 | 0 | 0 | 1 | 1 |
| 5111 | Farmers | manual | manual | 1 | 0 | 1 | 2 | 1 | 2 | 1 | 0 |
| 5112 | Horticultural trades | manual | manual | 1 | 0 | 1 | 1 | 1 | 2 | 1 | 0 |
| 5113 | Gardeners and landscape gardeners | manual | manual | 1 | 0 | 1 | 1 | 1 | 2 | 1 | 1 |
| 5114 | Groundsmen and greenkeepers | manual | manual | 1 | 0 | 1 | 1 | 1 | 2 | 1 | 0 |
| 5119 | Agricultural and fishing trades n.e.c. | manual | manual | 1 | 0 | 1 | 1 | 1 | 2 | 1 | 1 |
| 5211 | Smiths and forge workers | manual | manual | 1 | 1 | 1 | 3 | 2 | 2 | 1 | 1 |
| 5212 | Moulders, core makers and die casters | manual | manual | 2 | 1 | 1 | 3 | 1 | 2 | 1 | 1 |
| 5213 | Sheet metal workers | manual | manual | 2 | 1 | 1 | 3 | 2 | 2 | 1 | 1 |
| 5214 | Metal plate workers, and riveters | manual | manual | 2 | 1 | 1 | 1 | 2 | 2 | 1 | 1 |
| 5215 | Welding trades | manual | manual | 2 | 1 | 1 | 3 | 1 | 2 | 1 | 0 |
| 5216 | Pipe fitters | manual | manual | 1 | 2 | 2 | 3 | 2 | 2 | 1 | 1 |
| 5221 | Metal machining setters and setter-operators | manual | manual | 2 | 1 | 1 | 3 | 1 | 2 | 1 | 0 |
| 5222 | Tool makers, tool fitters and markers-out | manual | manual | 1 | 1 | 1 | 3 | 1 | 2 | 1 | 1 |
| 5223 | Metal working production and maintenance fitters | manual | manual | 1 | 1 | 1 | 2 | 2 | 2 | 1 | 1 |
| 5224 | Precision instrument makers and repairers | manual | manual | 1 | 0 | 1 | 3 | 1 | 2 | 1 | 1 |
| 5225 | Air-conditioning and refrigeration engineers | manual | manual | 1 | 2 | 2 | 3 | 2 | 2 | 1 | 1 |
| 5231 | Vehicle technicians, mechanics and electricians | manual | manual | 2 | 2 | 2 | 3 | 2 | 2 | 1 | 0 |
| 5232 | Vehicle body builders and repairers | manual | manual | 2 | 2 | 2 | 3 | 2 | 2 | 1 | 0 |
| 5234 | Vehicle paint technicians | manual | manual | 1 | 0 | 1 | 3 | 1 | 2 | 1 | 0 |
| 5235 | Aircraft maintenance and related trades | manual | manual | 2 | 1 | 1 | 3 | 2 | 2 | 1 | 0 |
| 5236 | Boat and ship builders and repairers | manual | manual | 1 | 1 | 1 | 2 | 2 | 2 | 1 | 0 |
| 5237 | Rail and rolling stock builders and repairers | manual | manual | 1 | 1 | 1 | 2 | 2 | 2 | 1 | 0 |
| 5241 | Electricians and electrical fitters | other workers | other workers-non-office based | 1 | 2 | 2 | 3 | 2 | 2 | 1 | 0 |
| 5242 | Telecommunications engineers | other workers | other workers-non-office based | 1 | 1 | 1 | 3 | 1 | 2 | 1 | 0 |
| 5244 | TV, video and audio engineers | other workers | other workers-non-office based | 1 | 2 | 2 | 3 | 2 | 2 | 1 | 0 |
| 5245 | IT engineers | other workers | other workers-non-office based | 1 | 0 | 1 | 3 | 1 | 2 | 1 | 0 |
| 5249 | Electrical and electronic trades n.e.c. | other workers | other workers-non-office based | 1 | 1 | 1 | 3 | 1 | 2 | 1 | 1 |
| 5250 | Skilled metal, electrical and electronic trades supervisors | manual | manual | 3 | 1 | 1 | 3 | 1 | 2 | 1 | 0 |
| 5311 | Steel erectors | manual | manual | 2 | 1 | 1 | 1 | 2 | 2 | 2 | 1 |
| 5312 | Bricklayers and masons | manual | manual | 1 | 1 | 1 | 1 | 2 | 2 | 2 | 1 |
| 5313 | Roofers, roof tilers and slaters | manual | manual | 1 | 1 | 1 | 1 | 2 | 2 | 2 | 0 |
| 5314 | Plumbers and heating and ventilating engineers | manual | manual | 1 | 2 | 2 | 3 | 2 | 2 | 2 | 1 |
| 5315 | Carpenters and joiners | manual | manual | 1 | 2 | 2 | 2 | 2 | 2 | 2 | 0 |
| 5316 | Glaziers, window fabricators and fitters | manual | manual | 1 | 2 | 2 | 2 | 2 | 2 | 2 | 0 |
| 5319 | Construction and building trades n.e.c. | manual | manual | 2 | 1 | 1 | 2 | 2 | 2 | 2 | 1 |
| 5321 | Plasterers | manual | manual | 1 | 2 | 2 | 3 | 2 | 2 | 1 | 0 |
| 5322 | Floorers and wall tilers | manual | manual | 1 | 2 | 2 | 3 | 2 | 2 | 1 | 0 |
| 5323 | Painters and decorators | manual | manual | 1 | 2 | 2 | 2 | 1 | 2 | 1 | 1 |
| 5330 | Construction and building trades supervisors | manual | manual | 3 | 1 | 1 | 2 | 1 | 2 | 3 | 1 |
| 5411 | Weavers and knitters | manual | manual | 1 | 0 | 1 | 3 | 1 | 2 | 1 | 0 |
| 5412 | Upholsterers | manual | manual | 1 | 1 | 1 | 3 | 1 | 2 | 1 | 0 |
| 5413 | Footwear and leather working trades | manual | manual | 1 | 0 | 2 | 3 | 1 | 2 | 1 | 0 |
| 5414 | Tailors and dressmakers | manual | manual | 1 | 2 | 2 | 3 | 2 | 2 | 1 | 0 |
| 5419 | Textiles, garments and related trades n.e.c. | manual | manual | 1 | 1 | 1 | 3 | 1 | 2 | 1 | 0 |
| 5421 | Pre-press technicians | manual | manual | 2 | 1 | 1 | 3 | 1 | 2 | 2 | 0 |
| 5422 | Printers | manual | manual | 2 | 1 | 1 | 3 | 2 | 2 | 2 | 0 |
| 5423 | Print finishing and binding workers | manual | manual | 2 | 1 | 1 | 3 | 1 | 2 | 2 | 0 |
| 5431 | Butchers | food processing | food processing | 3 | 2 | 2 | 3 | 2 | 2 | 2 | 0 |
| 5432 | Bakers and flour confectioners | food processing | food processing | 3 | 2 | 2 | 3 | 2 | 2 | 2 | 0 |
| 5433 | Fishmongers and poultry dressers | food processing | food processing | 3 | 2 | 2 | 3 | 2 | 2 | 2 | 1 |
| 5434 | Chefs | hospitality | hospitality | 1 | 1 | 1 | 3 | 2 | 2 | 2 | 1 |
| 5435 | Cooks | hospitality | hospitality | 1 | 1 | 1 | 3 | 2 | 2 | 2 | 1 |
| 5436 | Catering and bar managers | hospitality | hospitality | 2 | 3 | 2 | 3 | 1 | 2 | 2 | 1 |
| 5441 | Glass and ceramics makers, decorators and finishers | manual | manual | 1 | 1 | 1 | 3 | 1 | 2 | 2 | 0 |
| 5442 | Furniture makers and other craft woodworkers | manual | manual | 1 | 1 | 1 | 3 | 2 | 2 | 2 | 0 |
| 5443 | Florists | retail | retail | 3 | 3 | 2 | 3 | 1 | 2 | 2 | 0 |
| 5449 | Other skilled trades n.e.c. | manual | manual | 1 | 0 | 1 | 3 | 1 | 2 | 2 | 0 |
| 6121 | Nursery nurses and assistants | social and education | education | 3 | 3 | 2 | 3 | 3 | 2 | 1 | 1 |
| 6122 | Childminders and related occupations | social and education | education | 3 | 3 | 2 | 3 | 3 | 2 | 1 | 1 |
| 6123 | Playworkers | social and education | education | 3 | 3 | 2 | 3 | 3 | 2 | 1 | 1 |
| 6125 | Teaching assistants | social and education | education | 3 | 3 | 2 | 3 | 3 | 2 | 1 | 0 |
| 6126 | Educational support assistants | social and education | education | 3 | 3 | 2 | 3 | 3 | 2 | 1 | 1 |
| 6131 | Veterinary nurses | other workers | other workers-non-office based | 2 | 3 | 2 | 3 | 2 | 1 | 2 | 0 |
| 6132 | Pest control officers | sanitation services | sanitation services | 1 | 0 | 1 | 3 | 1 | 2 | 2 | 1 |
| 6139 | Animal care services occupations n.e.c. | other workers | other workers-non-office based | 1 | 2 | 2 | 3 | 1 | 2 | 2 | 1 |
| 6141 | Nursing auxiliaries and assistants | healthcare | healthcare-patient contact | 3 | 3 | 3 | 3 | 3 | 1 | 2 | 1 |
| 6142 | Ambulance staff (excluding paramedics) | healthcare | healthcare-patient contact | 2 | 3 | 3 | 2 | 3 | 1 | 2 | 2 |
| 6143 | Dental nurses | healthcare | healthcare-patient contact | 2 | 3 | 2 | 3 | 3 | 1 | 2 | 0 |
| 6144 | Houseparents and residential wardens | social and education | social care | 3 | 3 | 2 | 3 | 3 | 1 | 2 | 1 |
| 6145 | Care workers and home carers | social and education | social care | 2 | 3 | 2 | 3 | 3 | 1 | 2 | 2 |
| 6146 | Senior care workers | social and education | social care | 2 | 3 | 2 | 3 | 3 | 1 | 2 | 1 |
| 6147 | Care escorts | social and education | social care | 2 | 3 | 2 | 3 | 3 | 1 | 2 | 2 |
| 6148 | Undertakers, mortuary and crematorium assistants | social and education | social care | 2 | 3 | 2 | 3 | 1 | 2 | 2 | 1 |
| 6211 | Sports and leisure assistants | other workers | other workers-non-office based | 2 | 3 | 2 | 2 | 2 | 2 | 1 | 2 |
| 6212 | Travel agents | other workers | other workers-office based | 3 | 3 | 2 | 3 | 1 | 2 | 1 | 0 |
| 6214 | Air travel assistants | transport | transport-public facing | 3 | 3 | 2 | 3 | 3 | 1 | 1 | 1 |
| 6215 | Rail travel assistants | transport | transport-public facing | 3 | 3 | 2 | 3 | 2 | 1 | 1 | 1 |
| 6219 | Leisure and travel service occupations n.e.c. | transport | transport-public facing | 3 | 3 | 2 | 2 | 2 | 1 | 1 | 1 |
| 6221 | Hairdressers and barbers | personal care | personal care | 2 | 3 | 2 | 3 | 3 | 1 | 2 | 1 |
| 6222 | Beauticians and related occupations | personal care | personal care | 2 | 3 | 2 | 3 | 3 | 1 | 2 | 1 |
| 6231 | Housekeepers and related occupations | hospitality | hospitality | 1 | 2 | 2 | 3 | 2 | 2 | 2 | 0 |
| 6232 | Caretakers | hospitality | hospitality | 3 | 3 | 2 | 3 | 1 | 2 | 2 | 1 |
| 6240 | Cleaning and housekeeping managers and supervisors | hospitality | hospitality | 3 | 3 | 2 | 3 | 1 | 2 | 3 | 1 |
| 7111 | Sales and retail assistants | retail | retail | 3 | 3 | 2 | 3 | 2 | 1 | 1 | 1 |
| 7112 | Retail cashiers and check-out operators | retail | retail | 3 | 3 | 2 | 3 | 2 | 1 | 1 | 0 |
| 7113 | Telephone salespersons | other workers | other workers-office based | 3 | 0 | 1 | 3 | 1 | 2 | 1 | 0 |
| 7114 | Pharmacy and other dispensing assistants | healthcare | healthcare-patient contact | 3 | 3 | 3 | 3 | 1 | 1 | 1 | 1 |
| 7115 | Vehicle and parts salespersons and advisers | retail | retail | 3 | 3 | 2 | 3 | 2 | 1 | 1 | 0 |
| 7121 | Collector salespersons and credit agents | other workers | other workers-office based | 3 | 3 | 2 | 1 | 2 | 2 | 2 | 0 |
| 7122 | Debt, rent and other cash collectors | other workers | other workers-office based | 2 | 2 | 2 | 3 | 1 | 1 | 2 | 0 |
| 7123 | Roundspersons and van salespersons | other workers | other workers-non-office based | 3 | 3 | 2 | 1 | 2 | 2 | 2 | 0 |
| 7124 | Market and street traders and assistants | retail | retail | 3 | 3 | 2 | 1 | 2 | 2 | 2 | 0 |
| 7125 | Merchandisers and window dressers | other workers | other workers-non-office based | 2 | 3 | 2 | 3 | 1 | 2 | 2 | 0 |
| 7129 | Sales related occupations n.e.c. | retail | retail | 2 | 3 | 2 | 3 | 1 | 2 | 2 | 1 |
| 7130 | Sales supervisors | retail | retail | 3 | 3 | 2 | 3 | 1 | 1 | 1 | 1 |
| 7211 | Call and contact centre occupations | other workers | other workers-office based | 3 | 1 | 1 | 3 | 1 | 2 | 2 | 1 |
| 7213 | Telephonists | other workers | other workers-office based | 0 | 0 | 0 | 0 | 0 | 0 | 2 | 1 |
| 7214 | Communication operators | other workers | other workers-office based | 0 | 0 | 0 | 0 | 0 | 0 | 2 | 1 |
| 7215 | Market research interviewers | other workers | other workers-office based | 0 | 0 | 0 | 0 | 0 | 0 | 2 | 1 |
| 7219 | Customer service occupations n.e.c. | other workers | other workers-office based | 0 | 0 | 0 | 0 | 0 | 0 | 2 | 1 |
| 7220 | Customer service managers and supervisors | other workers | other workers-office based | 2 | 0 | 1 | 3 | 1 | 2 | 1 | 1 |
| 8111 | Food, drink and tobacco process operatives | food processing | food processing | 3 | 1 | 1 | 3 | 2 | 2 | 2 | 1 |
| 8112 | Glass and ceramics process operatives | manual | manual | 3 | 1 | 1 | 3 | 2 | 2 | 2 | 1 |
| 8113 | Textile process operatives | manual | manual | 3 | 1 | 1 | 3 | 2 | 2 | 2 | 1 |
| 8114 | Chemical and related process operatives | manual | manual | 3 | 1 | 1 | 3 | 2 | 2 | 2 | 1 |
| 8115 | Rubber process operatives | manual | manual | 3 | 1 | 1 | 3 | 2 | 2 | 2 | 1 |
| 8116 | Plastics process operatives | manual | manual | 3 | 1 | 1 | 3 | 2 | 2 | 2 | 0 |
| 8117 | Metal making and treating process operatives | manual | manual | 3 | 1 | 1 | 3 | 2 | 2 | 2 | 1 |
| 8118 | Electroplaters | manual | manual | 3 | 1 | 1 | 3 | 2 | 2 | 2 | 1 |
| 8119 | Process operatives n.e.c. | manual | manual | 3 | 1 | 1 | 2 | 2 | 2 | 2 | 1 |
| 8121 | Paper and wood machine operatives | manual | manual | 2 | 1 | 1 | 3 | 1 | 2 | 2 | 0 |
| 8122 | Coal mine operatives | manual | manual | 3 | 1 | 1 | 2 | 2 | 2 | 2 | 0 |
| 8123 | Quarry workers and related operatives | manual | manual | 3 | 1 | 1 | 2 | 2 | 2 | 2 | 0 |
| 8124 | Energy plant operatives | manual | manual | 2 | 1 | 1 | 3 | 1 | 2 | 2 | 0 |
| 8125 | Metal working machine operatives | manual | manual | 2 | 1 | 1 | 3 | 1 | 2 | 2 | 0 |
| 8126 | Water and sewerage plant operatives | manual | manual | 2 | 1 | 1 | 3 | 1 | 2 | 2 | 0 |
| 8127 | Printing machine assistants | manual | manual | 2 | 1 | 1 | 3 | 2 | 2 | 2 | 0 |
| 8129 | Plant and machine operatives n.e.c. | manual | manual | 3 | 1 | 1 | 3 | 2 | 2 | 2 | 0 |
| 8131 | Assemblers (electrical and electronic products) | manual | manual | 3 | 1 | 1 | 3 | 1 | 2 | 2 | 1 |
| 8132 | Assemblers (vehicles and metal goods) | manual | manual | 3 | 1 | 1 | 3 | 2 | 2 | 2 | 1 |
| 8133 | Routine inspectors and testers | manual | manual | 1 | 1 | 1 | 3 | 1 | 2 | 2 | 1 |
| 8134 | Weighers, graders and sorters | manual | manual | 1 | 1 | 1 | 3 | 1 | 2 | 2 | 1 |
| 8135 | Tyre, exhaust and windscreen fitters | manual | manual | 2 | 2 | 2 | 3 | 2 | 2 | 2 | 1 |
| 8137 | Sewing machinists | manual | manual | 3 | 1 | 1 | 3 | 2 | 2 | 2 | 1 |
| 8139 | Assemblers and routine operatives n.e.c. | manual | manual | 3 | 1 | 1 | 3 | 1 | 2 | 2 | 0 |
| 8141 | Scaffolders, stagers and riggers | manual | manual | 2 | 1 | 1 | 2 | 2 | 2 | 2 | 1 |
| 8142 | Road construction operatives | manual | manual | 3 | 1 | 1 | 1 | 2 | 2 | 2 | 1 |
| 8143 | Rail construction and maintenance operatives | manual | manual | 2 | 1 | 1 | 1 | 2 | 2 | 2 | 1 |
| 8149 | Construction operatives n.e.c. | manual | manual | 2 | 2 | 2 | 2 | 1 | 2 | 2 | 1 |
| 8211 | Large goods vehicle drivers | transport | transport-nonpublic facing | 1 | 0 | 0 | 3 | 1 | 2 | 2 | 1 |
| 8212 | Van drivers | transport | transport-nonpublic facing | 3 | 1 | 2 | 1 | 2 | 2 | 2 | 1 |
| 8213 | Bus and coach drivers | transport | transport-public facing | 3 | 3 | 2 | 3 | 2 | 2 | 2 | 1 |
| 8214 | Taxi and cab drivers and chauffeurs | transport | transport-public facing | 3 | 3 | 2 | 3 | 2 | 2 | 2 | 1 |
| 8215 | Driving instructors | transport | transport-public facing | 2 | 2 | 2 | 3 | 3 | 1 | 2 | 0 |
| 8221 | Crane drivers | other workers | other workers-non-office based | 1 | 0 | 0 | 2 | 1 | 2 | 2 | 1 |
| 8222 | Fork-lift truck drivers | other workers | other workers-non-office based | 1 | 1 | 1 | 1 | 1 | 2 | 2 | 1 |
| 8223 | Agricultural machinery drivers | other workers | other workers-non-office based | 1 | 0 | 1 | 2 | 1 | 2 | 2 | 1 |
| 8229 | Mobile machine drivers and operatives n.e.c. | other workers | other workers-non-office based | 1 | 0 | 1 | 2 | 1 | 2 | 2 | 1 |
| 8231 | Train and tram drivers | transport | transport-nonpublic facing | 1 | 0 | 1 | 3 | 2 | 2 | 1 | 0 |
| 8232 | Marine and waterways transport operatives | transport | transport-nonpublic facing | 2 | 1 | 1 | 1 | 2 | 2 | 1 | 1 |
| 8233 | Air transport operatives | transport | transport-nonpublic facing | 2 | 2 | 1 | 1 | 2 | 2 | 1 | 1 |
| 8234 | Rail transport operatives | transport | transport-nonpublic facing | 1 | 0 | 1 | 3 | 2 | 2 | 1 | 1 |
| 8239 | Other drivers and transport operatives n.e.c. | transport | transport-nonpublic facing | 2 | 1 | 1 | 2 | 1 | 2 | 1 | 2 |
| 9111 | Farm workers | manual | manual | 1 | 0 | 1 | 1 | 1 | 2 | 2 | 1 |
| 9112 | Forestry workers | manual | manual | 1 | 0 | 1 | 1 | 1 | 2 | 2 | 1 |
| 9119 | Fishing and other elementary agriculture occupations n.e.c. | manual | manual | 1 | 0 | 1 | 1 | 1 | 2 | 2 | 2 |
| 9120 | Elementary construction occupations | manual | manual | 3 | 1 | 1 | 1 | 2 | 2 | 1 | 0 |
| 9132 | Industrial cleaning process occupations | sanitation services | sanitation services | 3 | 1 | 1 | 3 | 2 | 2 | 3 | 1 |
| 9134 | Packers, bottlers, canners and fillers | manual | manual | 3 | 1 | 1 | 3 | 1 | 2 | 3 | 1 |
| 9139 | Elementary process plant occupations n.e.c. | manual | manual | 3 | 1 | 1 | 3 | 2 | 2 | 3 | 1 |
| 9211 | Postal workers, mail sorters, messengers and couriers | transport | transport-nonpublic facing | 3 | 3 | 2 | 3 | 1 | 2 | 1 | 1 |
| 9219 | Elementary administration occupations n.e.c. | other workers | other workers-office based | 2 | 1 | 1 | 3 | 1 | 2 | 1 | 1 |
| 9231 | Window cleaners | sanitation services | sanitation services | 1 | 0 | 1 | 2 | 1 | 2 | 2 | 0 |
| 9232 | Street cleaners | sanitation services | sanitation services | 1 | 1 | 1 | 1 | 1 | 2 | 2 | 1 |
| 9233 | Cleaners and domestics | sanitation services | sanitation services | 2 | 3 | 2 | 3 | 1 | 2 | 2 | 1 |
| 9234 | Launderers, dry cleaners and pressers | sanitation services | sanitation services | 1 | 3 | 2 | 3 | 1 | 2 | 2 | 0 |
| 9235 | Refuse and salvage occupations | sanitation services | sanitation services | 1 | 3 | 1 | 1 | 1 | 2 | 2 | 0 |
| 9236 | Vehicle valeters and cleaners | sanitation services | sanitation services | 1 | 3 | 2 | 1 | 1 | 2 | 2 | 0 |
| 9239 | Elementary cleaning occupations n.e.c. | sanitation services | sanitation services | 2 | 3 | 2 | 3 | 1 | 2 | 2 | 1 |
| 9241 | Security guards and related occupations | police and protective services | police and protective services | 3 | 2 | 2 | 3 | 2 | 2 | 2 | 1 |
| 9242 | Parking and civil enforcement occupations | police and protective services | police and protective services | 0 | 0 | 0 | 0 | 0 | 0 | 2 | 1 |
| 9244 | School midday and crossing patrol occupations | other workers | other workers-non-office based | 3 | 3 | 2 | 2 | 2 | 2 | 2 | 0 |
| 9249 | Elementary security occupations n.e.c. | police and protective services | police and protective services | 3 | 2 | 2 | 3 | 2 | 2 | 2 | 1 |
| 9251 | Shelf fillers | retail | retail | 3 | 3 | 2 | 3 | 2 | 2 | 2 | 1 |
| 9259 | Elementary sales occupations n.e.c. | retail | retail | 3 | 3 | 2 | 2 | 1 | 2 | 2 | 1 |
| 9260 | Elementary storage occupations | manual | manual | 2 | 2 | 1 | 1 | 2 | 2 | 2 | 1 |
| 9271 | Hospital porters | healthcare | healthcare-patient contact | 3 | 3 | 3 | 3 | 3 | 1 | 2 | 0 |
| 9272 | Kitchen and catering assistants | food processing | food processing | 1 | 3 | 1 | 3 | 2 | 2 | 2 | 2 |
| 9273 | Waiters and waitresses | hospitality | hospitality | 3 | 3 | 2 | 3 | 3 | 1 | 2 | 3 |
| 9274 | Bar staff | hospitality | hospitality | 3 | 3 | 2 | 3 | 2 | 1 | 2 | 3 |
| 9275 | Leisure and theme park attendants | hospitality | hospitality | 0 | 0 | 0 | 0 | 0 | 0 | 2 | 2 |
| 9279 | Other elementary services occupations n.e.c. | hospitality | hospitality | 3 | 3 | 2 | 3 | 1 | 2 | 2 | 1 |

Table S2: Outcome Missingness patterns of all covariates used for adjustment purposes

|  | **Self-Reported Long-COVID** | | | | |
| --- | --- | --- | --- | --- | --- |
| **Covariates** |  |  |  |  |  |
| **Age categories** | **No** | **Yes** | **Total (missing excluded)** | **Missing** | **Total** |
| 15-19 | 21,187 | 1,451 | 22,638 | 64 | 22,702 |
| % | 7.10 | 4.67 | 6.87 | 6.77 | 6.87 |
| 20-24 | 16,089 | 1,158 | 17,247 | 142 | 17,389 |
| % | 5.39 | 3.73 | 5.24 | 15.01 | 5.26 |
| 25-29 | 20,552 | 1,547 | 22,099 | 118 | 22,217 |
| % | 6.89 | 4.98 | 6.71 | 12.47 | 6.73 |
| 30-34 | 25,424 | 2,212 | 27,636 | 129 | 27,765 |
| % | 8.52 | 7.12 | 8.39 | 13.64 | 8.41 |
| 35-39 | 28,213 | 2,965 | 31,178 | 74 | 31,252 |
| % | 9.46 | 9.55 | 9.47 | 7.82 | 9.46 |
| 40-44 | 29,884 | 3,832 | 33,716 | 83 | 33,799 |
| % | 10.02 | 12.34 | 10.24 | 8.77 | 10.23 |
| 45-49 | 31,661 | 4,110 | 35,771 | 61 | 35,832 |
| % | 10.61 | 13.24 | 10.86 | 6.45 | 10.85 |
| 50-54 | 35,524 | 4,636 | 40,160 | 99 | 40,259 |
| % | 11.91 | 14.93 | 12.19 | 10.47 | 12.19 |
| 55-59 | 38,842 | 4,410 | 43,252 | 82 | 43,334 |
| % | 13.02 | 14.20 | 13.13 | 8.67 | 13.12 |
| 60-64 | 40,300 | 3,869 | 44,169 | 73 | 44,242 |
| % | 13.51 | 12.46 | 13.41 | 7.72 | 13.4 |
| 65-69 | 10,615 | 858 | 11,473 | 21 | 11,494 |
| % | 3.56 | 2.76 | 3.48 | 2.22 | 3.48 |
| **Total** | **298,291** | **31,048** | **329,339** | **946** | **330,285** |
| **%** | **100.00** | **100.00** | **100.00** | **100.00** | **100** |
| **Sex** |  |  |  |  |  |
| Male | 137,895 | 11,998 | 149,893 | 470 | 150,363 |
| % | 46.23 | 38.64 | 45.51 | 49.68 | 45.53 |
| Female | 160,396 | 19,050 | 179,446 | 476 | 179,922 |
| % | 53.77 | 61.36 | 54.49 | 50.32 | 54.47 |
| **Total** | **298,291** | **31,048** | **329,339** | **946** | **330,285** |
| **%** | **100** | **100** | **100** | **100** | **100** |
| **Ethnicity** |  |  |  |  |  |
| White | 272,298 | 28,655 | 300,953 | 833 | 301,786 |
| % | 91.29 | 92.29 | 91.38 | 88.05 | 91.37 |
| Mixed | 5,041 | 508 | 5,549 | 27 | 5,576 |
| % | 1.69 | 1.64 | 1.68 | 2.85 | 1.69 |
| Asian | 14,408 | 1,266 | 15,674 | 44 | 15,718 |
| % | 4.83 | 4.08 | 4.76 | 4.65 | 4.76 |
| Black | 3,533 | 310 | 3,843 | 22 | 3,865 |
| % | 1.18 | 1 | 1.17 | 2.33 | 1.17 |
| Other | 3,009 | 309 | 3,318 | 16 | 3,334 |
| % | 1.01 | 1 | 1.01 | 1.69 | 1.01 |
| . | 2 | 0 | 2 | 4 | 6 |
| % | 0 | 0 | 0.00 | 0.42 | 0 |
| **Total** | **298,291** | **31,048** | **329,339** | **946** | **330,285** |
| **%** | **100** | **100** |  | **100** | **100** |
| **Deprivation (Quintiles)** |  |  |  |  |  |
| 1st Quartile | 45,474 | 5,762 | 51,236 | 149 | 51,385 |
| % | 15.24 | 18.56 | 15.56 | 15.75 | 15.56 |
| 2nd Quartile | 69,967 | 7,477 | 77,444 | 268 | 77,712 |
| % | 23.46 | 24.08 | 23.51 | 28.33 | 23.53 |
| 3rd Quartile | 85,177 | 8,540 | 93,717 | 250 | 93,967 |
| % | 28.56 | 27.51 | 28.46 | 26.43 | 28.45 |
| 4th Quartile | 97,673 | 9,269 | 106,942 | 279 | 107,221 |
| % | 32.74 | 29.85 | 32.47 | 29.49 | 32.46 |
| **Total** | **298,291** | **31,048** | **329,339** | **946** | **330,285** |
| **%** | **100** | **100** |  | **100** | **100** |
| **UK Region** |  |  |  |  |  |
| North East | 10,169 | 1,368 | 11,537 | 18 | 11,555 |
| % | 3.41 | 4.41 | 3.50 | 1.9 | 3.5 |
| North West | 32,607 | 3,974 | 36,581 | 66 | 36,647 |
| % | 10.93 | 12.8 | 11.11 | 6.98 | 11.1 |
| Yorkshire & the Humber | 23,436 | 2,739 | 26,175 | 20 | 26,195 |
| % | 7.86 | 8.82 | 7.95 | 2.11 | 7.93 |
| East Midlands | 18,841 | 2,087 | 20,928 | 25 | 20,953 |
| % | 6.32 | 6.72 | 6.35 | 2.64 | 6.34 |
| West Midlands | 21,628 | 2,472 | 24,100 | 38 | 24,138 |
| % | 7.25 | 7.96 | 7.32 | 4.02 | 7.31 |
| East of England | 27,113 | 2,763 | 29,876 | 59 | 29,935 |
| % | 9.09 | 8.9 | 9.07 | 6.24 | 9.06 |
| London | 57,753 | 5,277 | 63,030 | 278 | 63,308 |
| % | 19.36 | 17 | 19.14 | 29.39 | 19.17 |
| South East | 36,803 | 3,532 | 40,335 | 78 | 40,413 |
| % | 12.34 | 11.38 | 12.25 | 8.25 | 12.24 |
| South West | 22,688 | 2,187 | 24,875 | 61 | 24,936 |
| % | 7.61 | 7.04 | 7.55 | 6.45 | 7.55 |
| Northern Ireland | 8,460 | 848 | 9,308 | 31 | 9,339 |
| % | 2.84 | 2.73 | 2.83 | 3.28 | 2.83 |
| Scotland | 24,275 | 2,280 | 26,555 | 120 | 26,675 |
| % | 8.14 | 7.34 | 8.06 | 12.68 | 8.08 |
| Wales | 14,518 | 1,521 | 16,039 | 152 | 16,191 |
| % | 4.87 | 4.9 | 4.87 | 16.07 | 4.9 |
| **Total** | **298,291** | **31,048** | **329,339** | **946** | **330,285** |
| **%** | **100** | **100** | **100.00** | **100** | **100** |
| **Urban or Rural** |  |  |  |  |  |
| Major urban | 112,006 | 11,720 | 123,726 | 389 | 124,115 |
| % | 37.55 | 37.75 | 37.57 | 41.12 | 37.58 |
| Urban city or town | 124,996 | 13,560 | 138,556 | 345 | 138,901 |
| % | 41.9 | 43.67 | 42.07 | 36.47 | 42.05 |
| Rural town | 29,599 | 2,938 | 32,537 | 97 | 32,634 |
| % | 9.92 | 9.46 | 9.88 | 10.25 | 9.88 |
| Rural village | 31,690 | 2,830 | 34,520 | 115 | 34,635 |
| % | 10.62 | 9.11 | 10.48 | 12.16 | 10.49 |
| **Total** | **298,291** | **31,048** | **329,339** | **946** | **330,285** |
| **%** | **100** | **100** | **100.00** | **100** | **100** |
| **Household Size** |  |  |  |  |  |
| 1 | 38,842 | 4,269 | 43,111 | 129 | 43,240 |
| % | 13.02 | 13.75 | 13.09 | 13.64 | 13.09 |
| 2 | 110,725 | 10,543 | 121,268 | 339 | 121,607 |
| % | 37.12 | 33.96 | 36.82 | 35.84 | 36.82 |
| 3 | 60,275 | 6,386 | 66,661 | 185 | 66,846 |
| % | 20.21 | 20.57 | 20.24 | 19.56 | 20.24 |
| 4 | 61,022 | 6,789 | 67,811 | 196 | 68,007 |
| % | 20.46 | 21.87 | 20.59 | 20.72 | 20.59 |
| 5+ | 27,427 | 3,061 | 30,488 | 97 | 30,585 |
| % | 9.19 | 9.86 | 9.26 | 10.25 | 9.26 |
| **Total** | **298,291** | **31,048** | **329,339** | **946** | **330,285** |
| **%** | **100** | **100** | **100.00** | **100** | **100** |
| **Health conditions** |  |  |  |  |  |
| No | 252,641 | 24,163 | 276,804 | 789 | 277,593 |
| % | 84.7 | 77.82 | 84.05 | 83.4 | 84.05 |
| Yes | 45,650 | 6,885 | 52,535 | 134 | 52,669 |
| % | 15.3 | 22.18 | 15.95 | 14.16 | 15.95 |
| . | 0 | 0 | 0 | 23 | 23 |
| % | **0** | **0** | **0.00** | **2.43** | **0.01** |
| **Total** | **298,291** | **31,048** | **329,339** | **946** | **330,285** |

Table S2: Outcome Missingness patterns of all exposures used

|  | **Self-Reported Long-COVID** | | | | |
| --- | --- | --- | --- | --- | --- |
| **Exposures** |  |  |  |  |  |
| **Industries (SIC)** | **No** | **Yes** | **Total (No missing)** | **Missing** | **Total** |
| Teaching and education | 25,568 | 3,708 | 29,276 | 60 | 29,336 |
| % | 8.72 | 12.14 | 9.04 | 6.56 | 9.04 |
| Health care | 22,569 | 2,762 | 25,331 | 71 | 25,402 |
| % | 7.70 | 9.04 | 7.82 | 7.76 | 7.82 |
| Social care | 5,959 | 845 | 6,804 | 22 | 6,826 |
| % | 2.03 | 2.77 | 2.10 | 2.40 | 2.10 |
| Transport (incl. storage, logistic) | 7,217 | 801 | 8,018 | 23 | 8,041 |
| % | 2.46 | 2.62 | 2.48 | 2.51 | 2.48 |
| Retail sector (incl.wholesale) | 12,669 | 1,413 | 14,082 | 41 | 14,123 |
| % | 4.32 | 4.63 | 4.35 | 4.48 | 4.35 |
| Hospitality (e.g. hotel restaurants) | 4,078 | 426 | 4,504 | 9 | 4,513 |
| % | 1.39 | 1.39 | 1.39 | 0.98 | 1.39 |
| Food production, agriculture, farming) | 3,517 | 349 | 3,866 | 16 | 3,882 |
| % | 1.20 | 1.14 | 1.19 | 1.75 | 1.20 |
| Personal services (e.g hairdressers) | 1,890 | 191 | 2,081 | 5 | 2,086 |
| % | 0.64 | 0.63 | 0.64 | 0.55 | 0.64 |
| Information technology and communication | 15,795 | 1,215 | 17,010 | 49 | 17,059 |
| % | 5.39 | 3.98 | 5.25 | 5.36 | 5.25 |
| Financial services inc. insurance | 17,761 | 1,424 | 19,185 | 52 | 19,237 |
| % | 6.06 | 4.66 | 5.93 | 5.68 | 5.92 |
| Manufacturing or construction | 20,206 | 2,034 | 22,240 | 55 | 22,295 |
| % | 6.89 | 6.66 | 6.87 | 6.01 | 6.87 |
| Civil service or Local Goverbment | 15,027 | 1,729 | 16,756 | 53 | 16,809 |
| % | 5.12 | 5.66 | 5.18 | 5.79 | 5.18 |
| Armed forces | 827 | 66 | 893 | 4 | 897 |
| % | 0.28 | 0.22 | 0.28 | 0.44 | 0.28 |
| Arts,Entertainment or Recreation | 4,530 | 416 | 4,946 | 19 | 4,965 |
| % | 1.54 | 1.36 | 1.53 | 2.08 | 1.53 |
| Other occupation sect | 30,576 | 2,816 | 33,392 | 113 | 33,505 |
| % | 10.43 | 9.22 | 10.31 | 12.35 | 10.32 |
| Not working | 105,034 | 10,348 | 115,382 | 323 | 115,705 |
| % | 35.82 | 33.88 | 35.64 | 35.30 | 35.64 |
| **Total** | **293,223** | **30,543** | **323,766** | **915** | **324,681** |
| **%** | **100** | **100.00** | **100.00** | **100.00** | **100.00** |
| **Occupations (Constructed by authors)** |  |  |  |  |  |
| education | 12,103 | 1,883 | 13,986 | 43 | 14,029 |
| % | 4.81 | 7.27 | 5.04 | 5.22 | 5.04 |
| food processing | 1,167 | 138 | 1,305 | 8 | 1,313 |
| % | 0.46 | 0.53 | 0.47 | 0.97 | 0.47 |
| healthcare-office based | 862 | 94 | 956 | 1 | 957 |
| % | 0.34 | 0.36 | 0.34 | 0.12 | 0.34 |
| healthcare-patient contact | 11,787 | 1,370 | 13,157 | 32 | 13,189 |
| % | 4.68 | 5.29 | 4.74 | 3.88 | 4.74 |
| Hospitality | 2,390 | 310 | 2,700 | 5 | 2,705 |
| % | 0.95 | 1.20 | 0.97 | 0.61 | 0.97 |
| Manual | 11,520 | 1,107 | 12,627 | 41 | 12,668 |
| % | 4.58 | 4.28 | 4.55 | 4.98 | 4.55 |
| other workers-non-office based | 12,160 | 1,151 | 13,311 | 39 | 13,350 |
| % | 4.83 | 4.45 | 4.80 | 4.73 | 4.80 |
| other workers-office based | 73,628 | 6,741 | 80,369 | 254 | 80,623 |
| % | 29.26 | 26.04 | 28.96 | 30.83 | 28.97 |
| personal care | 334 | 37 | 371 | 1 | 372 |
| % | 0.13 | 0.14 | 0.13 | 0.12 | 0.13 |
| police and protective services | 3,235 | 418 | 3,653 | 19 | 3,672 |
| % | 1.29 | 1.61 | 1.32 | 2.31 | 1.32 |
| Retail | 5,821 | 655 | 6,476 | 20 | 6,496 |
| % | 2.31 | 2.53 | 2.33 | 2.43 | 2.33 |
| sanitation services | 1,697 | 204 | 1,901 | 1 | 1,902 |
| % | 0.67 | 0.79 | 0.69 | 0.12 | 0.68 |
| social care | 7,006 | 1,093 | 8,099 | 27 | 8,126 |
| % | 2.78 | 4.22 | 2.92 | 3.28 | 2.92 |
| transport-nonpublic facing | 3,202 | 360 | 3,562 | 10 | 3,572 |
|  | 1.27 | 1.39 | 1.28 | 1.21 | 1.28 |
| transport-public facing | 970 | 118 | 1,088 | 1 | 1,089 |
| % | 0.39 | 0.46 | 0.39 | 0.12 | 0.39 |
| Not working | 103,717 | 10,209 | 113,926 | 322 | 114,248 |
| % | 41.22 | 39.44 | 41.06 | 39.08 | 41.05 |
| **Total** | **251,599** | **25,888** | **277,487** | **824** | **278,311** |
| **%** | **100** | **100** | **100** | **100** | **100** |
| **Occupations (SOC)** |  |  |  |  |  |
| Managers, directors and senior officials | 20,812 | 2,126 | 22,938 | 50 | 22,988 |
| % | 7.48 | 7.34 | 7.46 | 5.79 | 7.46 |
| Professional occupations | 54,576 | 5,476 | 60,052 | 170 | 60,222 |
| % | 19.60 | 18.90 | 19.54 | 19.70 | 19.54 |
| Associate professional and technical | 32,017 | 3,263 | 35,280 | 102 | 35,382 |
| % | 11.50 | 11.26 | 11.48 | 11.82 | 11.48 |
| Admin and secretarial | 22,839 | 2,347 | 25,186 | 59 | 25,245 |
| % | 8.20 | 8.10 | 8.19 | 6.84 | 8.19 |
| Skilled trades | 12,000 | 1,168 | 13,168 | 37 | 13,205 |
| % | 4.31 | 4.03 | 4.28 | 4.29 | 4.28 |
| Caring, leisure and other service | 11,425 | 1,956 | 13,381 | 52 | 13,433 |
| % | 4.10 | 6.75 | 4.35 | 6.03 | 4.36 |
| Sales and customer service | 7,805 | 934 | 8,739 | 27 | 8,766 |
| % | 2.80 | 3.22 | 2.84 | 3.13 | 2.84 |
| Process plant and machine operatives | 5,937 | 658 | 6,595 | 19 | 6,614 |
| % | 2.13 | 2.27 | 2.15 | 2.20 | 2.15 |
| Elementary | 7,255 | 843 | 8,098 | 25 | 8,123 |
| % | 2.61 | 2.91 | 2.63 | 2.90 | 2.64 |
| Not working | 103,717 | 10,209 | 113,926 | 322 | 114,248 |
| % | 37.26 | 35.23 | 37.07 | 37.31 | 37.07 |
| **Total** | **278,383** | **28,980** | **307,363** | **863** | **308,226** |
| **%** | **100.00** | **100.00** | **100.00** | **100.00** | **100.00** |

Table S3. Working and not working group by prevalence of health conditions (a: Long-Covid sample, b: reduced function sample)

| **Health condition** | | | | | | | | | |
| --- | --- | --- | --- | --- | --- | --- | --- | --- | --- |
| **(a)** | **Industry (SIC)** | | | **Occupation** | | | **SOC major group** | | |
| **Health conditions** | **Working** | **Non-working** | **TOTAL** | **Working** | **Non-working** | **TOTAL** | **Working** | **Non-working** | **TOTAL** |
| **No** | 183 265 | 88 516 | **271 781** | 143 161 | 87 238 | **230 399** | 169 865 | 87 238 | **257 103** |
| **%** | 87.95 | 76.72 | **83.94** | 87.53 | 76.57 | **83.03** | 87.81 | 76.57 | **83.65** |
| **Yes** | 25 119 | 26 866 | **51 985** | 20 400 | 26 688 | **47 088** | 23 572 | 26 688 | **50 260** |
| **%** | 12.05 | 23.28 | **16.06** | 12.47 | 23.43 | **16.97** | 12.19 | 23.43 | **16.35** |
| **TOTAL** | 208 384 | 115 382 | **323 766** | 163 561 | 113 926 | **277 487** | 193 437 | 113 926 | **307 363** |
| **%** | 64.36 | 35.64 | **100.00** | 58.94 | 41.06 | **100.00** | 62.93 | 37.07 | **100.00** |

| **Health condition** | | | | | | | | | |
| --- | --- | --- | --- | --- | --- | --- | --- | --- | --- |
| **(b)** | **Industry (SIC)** | | | **Occupation** | | | **SOC major group** | | |
| **Health conditions** | **Working** | **Non-working** | **TOTAL** | **Working** | **Non-working** | **TOTAL** | **Working** | **Non-working** | **TOTAL** |
| **No** | 16 702 | 7 025 | **23 727** | 12 894 | 6 913 | **19 807** | 15 508 | 6 913 | **22 421** |
|  | 82.70 | 67.89 | **77.68** | 82.24 | 67.71 | **76.51** | 82.62 | 67.71 | **77.37** |
| **Yes** | 3 493 | 3 323 | **6 816** | 2 785 | 3 296 | **6 081** | 3 262 | 3 296 | **6 558** |
|  | 17.30 | 32.11 | **22.32** | 17.76 | 32.29 | **23.49** | 17.38 | 32.29 | **22.63** |
| **TOTAL** | 20 195 | 10 348 | **30 543** | 15 679 | 10 209 | **25 888** | 18 770 | 10 209 | **28 979** |
|  | 100.00 | 100.00 | **100.00** | 100.00 | 100.00 | **100.00** | 100.00 | 100.00 | **100.00** |

Table S4. Working and not working group across industries by prevalence of long-COVID symptoms (a) and reduced function (b) and age sex and health conditions

**a.**

|  | **Exposure groups** | **Industry (SIC)** | | | | | | | | | | | | | | | | | | | |
| --- | --- | --- | --- | --- | --- | --- | --- | --- | --- | --- | --- | --- | --- | --- | --- | --- | --- | --- | --- | --- | --- |
|  |  | **Working** | | | | | | **Not working** | | | | | | **Total** | | | | | | | |
|  |  | **Count** | | | **%** | | | **Count** | | | **%** | | | **Count** | | | | **%** | | | |
| **Outcome** | **Self-reported long-COVID** | **No** | **Yes** | **Total** | **No** | **Yes** | **Total** | **No** | **Yes** | **Total** | **No** | **Yes** | **Total** | **No** | **Yes** | | **Total** | **No** | | **Yes** | **Total** |
| Age bands | 15-19 | 1 564 | 126 | 1690 | 92.5 | 7.5 | 100.0 | 19519 | 1314 | 20833 | 93.69 | 6.31 | 100.00 | 21083 | | 1440 | 22523 | | 93.6 | 6.4 | 100.0 |
|  | 20-24 | 7 944 | 605 | 8549 | 92.9 | 7.1 | 100.0 | 7876 | 535 | 8411 | 93.6 | 6.4 | 100.0 | 15820 | | 1140 | 16960 | | 93.3 | 6.7 | 100.0 |
|  | 25-29 | 15 880 | 1211 | 17091 | 92.9 | 7.1 | 100.0 | 4311 | 305 | 4616 | 93.4 | 6.6 | 100.0 | 20191 | | 1516 | 21707 | | 93.0 | 7.0 | 100.0 |
|  | 30-34 | 19 822 | 1654 | 21476 | 92.3 | 7.7 | 100.0 | 5136 | 514 | 5650 | 90.9 | 9.1 | 100.0 | 24958 | | 2168 | 27126 | | 92.0 | 8.0 | 100.0 |
|  | 35-39 | 22 124 | 2215 | 24339 | 90.9 | 9.1 | 100.0 | 5523 | 696 | 6219 | 88.8 | 11.2 | 100.0 | 27647 | | 2911 | 30558 | | 90.5 | 9.5 | 100.0 |
|  | 40-44 | 23 978 | 2965 | 26943 | 89.0 | 11.0 | 100.0 | 5246 | 801 | 6047 | 86.8 | 13.3 | 100.0 | 29224 | | 3766 | 32990 | | 88.6 | 11.4 | 100.0 |
|  | 45-49 | 25 282 | 3192 | 28474 | 88.8 | 11.2 | 100.0 | 5707 | 840 | 6547 | 87.2 | 12.8 | 100.0 | 30989 | | 4032 | 35021 | | 88.5 | 11.5 | 100.0 |
|  | 50-54 | 27 137 | 3421 | 30558 | 88.8 | 11.2 | 100.0 | 7672 | 1129 | 8801 | 87.2 | 12.8 | 100.0 | 34809 | | 4550 | 39359 | | 88.4 | 11.6 | 100.0 |
|  | 55-59 | 24 935 | 2818 | 27753 | 89.9 | 10.2 | 100.0 | 13245 | 1535 | 14780 | 89.6 | 10.4 | 100.0 | 38180 | | 4353 | 42533 | | 89.8 | 10.2 | 100.0 |
|  | 60-64 | 16 901 | 1780 | 18681 | 90.5 | 9.5 | 100.0 | 22918 | 2039 | 24957 | 91.8 | 8.2 | 100.0 | 39819 | | 3819 | 43638 | | 91.2 | 8.8 | 100.0 |
|  | 65-69 | 2 622 | 208 | 2830 | 92.7 | 7.4 | 100.0 | 7881 | 640 | 8521 | 92.5 | 7.5 | 100.0 | 10503 | | 848 | 11351 | | 92.5 | 7.5 | 100.0 |
| Sex | Male | 92 524 | 8323 | 100847 | 91.8 | 8.3 | 100.0 | 42782 | 3436 | 46218 | 92.6 | 7.4 | 100.0 | 135306 | | 11759 | 147065 | | 92.0 | 8.0 | 100.0 |
|  | Female | 95 665 | 11872 | 107537 | 89.0 | 11.0 | 100.0 | 62252 | 6912 | 69164 | 90.0 | 10.0 | 100.0 | 157917 | | 18784 | 176701 | | 89.4 | 10.6 | 100.0 |
| Health  Condition | No | 166 564 | 16701 | 183265 | 90.9 | 9.1 | 100.0 | 81490 | 7026 | 88516 | 92.1 | 7.9 | 100.0 | 248054 | | 23727 | 271781 | | 91.3 | 8.7 | 100.0 |
|  | Yes | 21 625 | 3494 | 25119 | 86.1 | 13.9 | 100.0 | 23544 | 3322 | 26866 | 87.6 | 12.4 | 100.0 | 45169 | | 6816 | 51985 | | 86.9 | 13.1 | 100.0 |
| **Total** |  | **188,189** | **20195** | **208384** | **90.3** | **9.7** | **100.0** | **105034** | **10348** | **115382** | **91.0** | **9.0** | **100.0** | **293223** | | **30543** | **323766** | | **90.6** | **9.4** | **100.0** |

**b.**

|  |  | **Working** | | | | | | | | **Not working** | | | | | | | |
| --- | --- | --- | --- | --- | --- | --- | --- | --- | --- | --- | --- | --- | --- | --- | --- | --- | --- |
|  |  | **Count** | | | | **%** | | | | **Count** | | | | **%** | | | |
| **Outcome** | **Reduced function** | **No** | **Yes, a little** | **Yes, a lot** | **Total** | **No** | **Yes, a little** | **Yes, a lot** | **Total** | **No** | **Yes, a little** | **Yes, a lot** | **Total** | **No** | **Yes, a little** | **Yes, a lot** | **Total** |
| Age bands | 15-19 | 51 | 55 | 20 | 126 | 40.48 | 43.65 | 15.87 | 100 | 536 | 593 | 184 | 1313 | 40.82 | 45.16 | 14.01 | 100 |
|  | 20-24 | 236 | 297 | 72 | 605 | 39.01 | 49.09 | 11.9 | 100 | 200 | 256 | 79 | 535 | 37.38 | 47.85 | 14.77 | 100 |
|  | 25-29 | 445 | 588 | 178 | 1211 | 36.75 | 48.55 | 14.7 | 100 | 96 | 133 | 76 | 305 | 31.48 | 43.61 | 24.92 | 100 |
|  | 30-34 | 530 | 834 | 290 | 1654 | 32.04 | 50.42 | 17.53 | 100 | 143 | 246 | 125 | 514 | 27.82 | 47.86 | 24.32 | 100 |
|  | 35-39 | 658 | 1122 | 435 | 2215 | 29.71 | 50.65 | 19.64 | 100 | 177 | 308 | 211 | 696 | 25.43 | 44.25 | 30.32 | 100 |
|  | 40-44 | 904 | 1468 | 593 | 2965 | 30.49 | 49.51 | 20 | 100 | 176 | 366 | 259 | 801 | 21.97 | 45.69 | 32.33 | 100 |
|  | 45-49 | 960 | 1574 | 658 | 3192 | 30.08 | 49.31 | 20.61 | 100 | 176 | 376 | 288 | 840 | 20.95 | 44.76 | 34.29 | 100 |
|  | 50-54 | 936 | 1678 | 808 | 3422 | 27.35 | 49.04 | 23.61 | 100 | 209 | 433 | 488 | 1130 | 18.50 | 38.32 | 43.19 | 100 |
|  | 55-59 | 753 | 1379 | 687 | 2819 | 26.71 | 48.92 | 24.37 | 100 | 291 | 672 | 572 | 1535 | 18.96 | 43.78 | 37.26 | 100 |
|  | 60-64 | 442 | 859 | 477 | 1778 | 24.86 | 48.31 | 26.83 | 100 | 459 | 907 | 673 | 2039 | 22.51 | 44.48 | 33.01 | 100 |
|  | 65-69 | 44 | 118 | 46 | 208 | 21.15 | 56.73 | 22.12 | 100 | 150 | 292 | 198 | 640 | 23.44 | 45.63 | 30.94 | 100 |
| Sex | Male | 2,670 | 4140 | 1513 | 8323 | 32.08 | 49.74 | 18.18 | 100 | 947 | 1498 | 991 | 3436 | 27.56 | 43.6 | 28.84 | 100 |
|  | Female | 3,289 | 5832 | 2751 | 11872 | 27.70 | 49.12 | 23.17 | 100 | 1,666 | 3084 | 2162 | 6912 | 24.10 | 44.62 | 31.28 | 100 |
| Health  Condition | No | 5,219 | 8317 | 3166 | 16702 | 31.25 | 49.8 | 18.96 | 100 | 2,152 | 3271 | 1602 | 7025 | 30.63 | 46.56 | 22.8 | 100 |
|  | Yes | 740 | 1655 | 1098 | 3493 | 29.51 | 49.38 | 31.43 | 100 | 461 | 1311 | 1551 | 3323 | 13.87 | 39.45 | 46.67 | 100 |
| **Total** |  | **5,959** | **9972** | **4264** | **20195** | **29.51** | **49.38** | **21.11** | **100** | **2,613** | **4582** | **3153** | **10348** | **25.25** | **44.28** | **30.47** | **100** |

Table S5. Working and not working group across occupations by prevalence of long-COVID symptoms and age, sex and health conditions

**a.**

|  | **Exposure groups** | **Occupations** | | | | | | | | | | | | | | | | | |
| --- | --- | --- | --- | --- | --- | --- | --- | --- | --- | --- | --- | --- | --- | --- | --- | --- | --- | --- | --- |
|  |  | **Working** | | | | | | **Not working** | | | | | | **Total** | | | | | |
|  |  | **Count** | | | **%** | | | **Count** | | | **%** | | | **Count** | | | **%** | | |
| **Outcome** | **Self-reported long-COVID** | **No** | **Yes** | **Total** | **No** | **Yes** | **Total** | **No** | **Yes** | **Total** | **No** | **Yes** | **Total** | **No** | **Yes** | **Total** | **No** | **Yes** | **Total** |
| Age bands | 15-19 | 1 325 | 107 | 1 432 | 92.5 | 7.5 | 100.0 | 19 446 | 1308 | 20 754 | 93.7 | 6.3 | 100.0 | 20 771 | 1415 | 22 186 | 93.6 | 6.4 | 100.0 |
|  | 20-24 | 6 385 | 469 | 6 854 | 93.2 | 6.8 | 100.0 | 7 764 | 529 | 8 293 | 93.6 | 6.4 | 100.0 | 14 149 | 998 | 15 147 | 93.4 | 6.6 | 100.0 |
|  | 25-29 | 12 418 | 935 | 13 353 | 93.0 | 7.0 | 100.0 | 4 189 | 299 | 4 488 | 93.3 | 6.7 | 100.0 | 16 607 | 1234 | 17 841 | 93.1 | 6.9 | 100.0 |
|  | 30-34 | 15 467 | 1273 | 16 740 | 92.4 | 7.6 | 100.0 | 5 025 | 507 | 5 532 | 90.8 | 9.2 | 100.0 | 20 492 | 1780 | 22 272 | 92.0 | 8.0 | 100.0 |
|  | 35-39 | 17 201 | 1665 | 18 866 | 91.2 | 8.8 | 100.0 | 5 395 | 683 | 6 078 | 88.8 | 11.2 | 100.0 | 22 596 | 2348 | 24 944 | 90.6 | 9.4 | 100.0 |
|  | 40-44 | 18 690 | 2299 | 20 989 | 89.1 | 11.0 | 100.0 | 5 097 | 766 | 5 863 | 86.9 | 13.1 | 100.0 | 23 787 | 3065 | 26 852 | 88.6 | 11.4 | 100.0 |
|  | 45-49 | 19 756 | 2499 | 22 255 | 88.8 | 11.2 | 100.0 | 5 562 | 824 | 6 386 | 87.1 | 12.9 | 100.0 | 25 318 | 3323 | 28 641 | 88.4 | 11.6 | 100.0 |
|  | 50-54 | 21 355 | 2639 | 23 994 | 89.0 | 11.0 | 100.0 | 7 511 | 1113 | 8 624 | 87.1 | 12.9 | 100.0 | 28 866 | 3752 | 32 618 | 88.5 | 11.5 | 100.0 |
|  | 55-59 | 19 711 | 2229 | 21 940 | 89.8 | 10.2 | 100.0 | 13 062 | 1515 | 14 577 | 89.6 | 10.4 | 100.0 | 32 773 | 3744 | 36 517 | 89.7 | 10.3 | 100.0 |
|  | 60-64 | 13 454 | 1409 | 14 863 | 90.5 | 9.5 | 100.0 | 22 807 | 2026 | 24 833 | 91.8 | 8.2 | 100.0 | 36 261 | 3435 | 39 696 | 91.3 | 8.7 | 100.0 |
|  | 65-69 | 2 120 | 155 | 2 275 | 93.2 | 6.8 | 100.0 | 7 859 | 639 | 8 498 | 92.5 | 7.5 | 100.0 | 9 979 | 794 | 10 773 | 92.6 | 7.4 | 100.0 |
| Sex | Male | 72 716 | 6444 | 79 160 | 91.9 | 8.1 | 100.0 | 42 226 | 3382 | 45 608 | 92.6 | 7.4 | 100.0 | 114 942 | 9826 | 124 768 | 92.1 | 7.9 | 100.0 |
|  | Female | 75 166 | 9235 | 84 401 | 89.1 | 10.9 | 100.0 | 61 491 | 6827 | 68 318 | 90.0 | 10.0 | 100.0 | 136 657 | 16062 | 152 719 | 89.5 | 10.5 | 100.0 |
| Health  Condition | No | 130 267 | 12894 | 143 161 | 91.0 | 9.0 | 100.0 | 80 324 | 6914 | 87 238 | 92.1 | 7.9 | 100.0 | 210 591 | 19808 | 230 399 | 91.4 | 8.6 | 100.0 |
|  | Yes | 17 615 | 2785 | 20 400 | 86.4 | 13.7 | 100.0 | 23 393 | 3295 | 26 688 | 87.7 | 12.4 | 100.0 | 41 008 | 6080 | 47 088 | 87.1 | 12.9 | 100.0 |
| **Total** |  | **147 882** | **15 679** | **163 561** | **90.4** | **9.6** | **100.0** | **103 717** | **10 209** | **113 926** | **91.0** | **9.0** | **100.0** | **251 599** | **25 888** | **277 487** | **90.7** | **9.3** | **100.0** |

**b.**

|  |  | **Working** | | | | | | | | | **Not working** | | | | | | | |
| --- | --- | --- | --- | --- | --- | --- | --- | --- | --- | --- | --- | --- | --- | --- | --- | --- | --- | --- |
|  |  | **Count** | | | | **%** | | | | **Count** | | | | | **%** | | | |
| **Outcome** | **Reduced function** | **No** | **Yes, a little** | **Yes, a lot** | **Total** | **No** | **Yes, a little** | **Yes, a lot** | **Total** | **No** | | **Yes, a little** | **Yes, a lot** | **Total** | **No** | **Yes, a little** | **Yes, a lot** | **Total** |
| Age bands | 15-19 | 47 | 46 | 14 | 107 | 43.93 | 42.99 | 13.08 | 100 | 535 | | 588 | 184 | 1 307 | 40.93 | 44.99 | 14.08 | 100 |
|  | 20-24 | 195 | 226 | 48 | 469 | 41.58 | 48.19 | 10.23 | 100 | 198 | | 255 | 76 | 529 | 37.43 | 48.2 | 14.37 | 100 |
|  | 25-29 | 334 | 462 | 139 | 935 | 35.72 | 49.41 | 14.87 | 100 | 95 | | 129 | 75 | 299 | 31.77 | 43.14 | 25.08 | 100 |
|  | 30-34 | 403 | 644 | 226 | 1 273 | 31.66 | 50.59 | 17.75 | 100 | 142 | | 243 | 122 | 507 | 28.01 | 47.93 | 24.06 | 100 |
|  | 35-39 | 509 | 843 | 313 | 1 665 | 30.57 | 50.63 | 18.8 | 100 | 173 | | 301 | 209 | 683 | 25.33 | 44.07 | 30.6 | 100 |
|  | 40-44 | 677 | 1 151 | 471 | 2 299 | 29.45 | 50.07 | 20.49 | 100 | 164 | | 353 | 249 | 766 | 21.41 | 46.08 | 32.51 | 100 |
|  | 45-49 | 752 | 1 252 | 494 | 2 498 | 30.10 | 50.12 | 19.78 | 100 | 171 | | 369 | 284 | 824 | 20.75 | 44.78 | 34.47 | 100 |
|  | 50-54 | 726 | 1 296 | 618 | 2 640 | 27.50 | 49.09 | 23.41 | 100 | 203 | | 427 | 484 | 1 114 | 18.22 | 38.33 | 43.45 | 100 |
|  | 55-59 | 623 | 1 056 | 551 | 2 230 | 27.94 | 47.35 | 24.71 | 100 | 283 | | 663 | 569 | 1 515 | 18.68 | 43.76 | 37.56 | 100 |
|  | 60-64 | 356 | 684 | 368 | 1 408 | 25.28 | 48.58 | 26.14 | 100 | 456 | | 901 | 669 | 2 026 | 22.51 | 44.47 | 33.02 | 100 |
|  | 65-69 | 31 | 84 | 40 | 155 | 20.00 | 54.19 | 25.81 | 100 | 150 | | 292 | 197 | 639 | 23.47 | 45.7 | 30.83 | 100 |
| Sex | Male | 2 081 | 3 189 | 1 173 | 6 443 | 32.30 | 49.50 | 18.21 | 100 | 929 | | 1474 | 979 | 3 382 | 27.47 | 43.58 | 28.95 | 100 |
|  | Female | 2 572 | 4 555 | 2 109 | 9 236 | 27.85 | 49.32 | 22.83 | 100 | 1 641 | | 3047 | 2139 | 6 827 | 24.04 | 44.63 | 31.33 | 100 |
| Health  Condition | No | 4 052 | 6 440 | 2 402 | 12 894 | 31.43 | 49.95 | 18.63 | 100 | 2 115 | | 3224 | 1574 | 6 913 | 30.59 | 46.64 | 22.77 | 100 |
|  | Yes | 601 | 1 304 | 880 | 2785 | 29.68 | 49.39 | 31.60 | 100 | 455 | | 1297 | 1544 | 3 296 | 13.80 | 39.35 | 46.84 | 100 |
| **Total** |  | **4,653** | **7744** | **3282** | **15679** | **29.68** | **49.39** | **20.93** | **100** | **2,570** | | **4521** | **3118** | **10209** | **25.17** | **44.28** | **30.54** | **100** |

Table S6. Working and not working group across major occupational groups by prevalence of long-COVID symptoms and age, sex and health conditions

**a.**

|  | **Exposure groups** | **Major occupational groups (SOC)** | | | | | | | | | | | | | | | | | |
| --- | --- | --- | --- | --- | --- | --- | --- | --- | --- | --- | --- | --- | --- | --- | --- | --- | --- | --- | --- |
|  |  | **Working** | | | | | | **Not working** | | | | | | **Total** | | | | | |
|  |  | **Count** | | | **%** | | | **Count** | | | **%** | | | **Count** | | | **%** | | |
| **Outcome** | **Self-reported long-COVID** | **No** | **Yes** | **Total** | **No** | **Yes** | **Total** | **No** | **Yes** | **Total** | **No** | **Yes** | **Total** | **No** | **Yes** | **Total** | **No** | **Yes** | **Total** |
| Age bands | 15-19 | 1 465 | 122 | 1 587 | 92.3 | 7.7 | 100.0 | 19 446 | 1 308 | 20 754 | 93.7 | 6.3 | 100.0 | 20 911 | 1 430 | 22 341 | 93.6 | 6.4 | 100.0 |
|  | 20-24 | 7 429 | 560 | 7 989 | 93.0 | 7.0 | 100.0 | 7 764 | 529 | 8 293 | 93.6 | 6.4 | 100.0 | 15 193 | 1 089 | 16 282 | 93.3 | 6.7 | 100.0 |
|  | 25-29 | 14 675 | 1 133 | 15 808 | 92.8 | 7.2 | 100.0 | 4 189 | 299 | 4 488 | 93.3 | 6.7 | 100.0 | 18 864 | 1 432 | 20 296 | 92.9 | 7.1 | 100.0 |
|  | 30-34 | 18 365 | 1 538 | 19 903 | 92.3 | 7.7 | 100.0 | 5 025 | 507 | 5 532 | 90.8 | 9.2 | 100.0 | 23 390 | 2 045 | 25 435 | 92.0 | 8.0 | 100.0 |
|  | 35-39 | 20 95 | 2 032 | 22 527 | 91.0 | 9.0 | 100.0 | 5 395 | 683 | 6 078 | 88.8 | 11.2 | 100.0 | 25 890 | 2 715 | 28 605 | 90.5 | 9.5 | 100.0 |
|  | 40-44 | 22 218 | 2 754 | 24 972 | 89.0 | 11.0 | 100.0 | 5 097 | 766 | 5 863 | 86.9 | 13.1 | 100.0 | 27 315 | 3 520 | 30 835 | 88.6 | 11.4 | 100.0 |
|  | 45-49 | 23 421 | 2 983 | 26 404 | 88.7 | 11.3 | 100.0 | 5 562 | 824 | 6 386 | 87.1 | 12.9 | 100.0 | 28 983 | 3 807 | 32 790 | 88.4 | 11.6 | 100.0 |
|  | 50-54 | 25 160 | 3 157 | 28 317 | 88.9 | 11.2 | 100.0 | 7 511 | 1 113 | 8 624 | 87.1 | 12.9 | 100.0 | 32 671 | 4 270 | 36 941 | 88.4 | 11.6 | 100.0 |
|  | 55-59 | 23 190 | 2 631 | 25 821 | 89.8 | 10.2 | 100.0 | 13 062 | 1 515 | 14 577 | 89.6 | 10.4 | 100.0 | 36 252 | 4 146 | 40 398 | 89.7 | 10.3 | 100.0 |
|  | 60-64 | 15 778 | 1 668 | 17 446 | 90.4 | 9.6 | 100.0 | 22 807 | 2 026 | 24 833 | 91.8 | 8.2 | 100.0 | 38 585 | 3 694 | 42 279 | 91.3 | 8.7 | 100.0 |
|  | 65-69 | 2 470 | 193 | 2 663 | 92.8 | 7.3 | 100.0 | 7 859 | 639 | 8 498 | 92.5 | 7.5 | 100.0 | 10 329 | 832 | 11 161 | 92.5 | 7.5 | 100.0 |
| Sex | Male | 85 553 | 7 693 | 93 246 | 91.8 | 8.3 | 100.0 | 42 226 | 3 382 | 45 608 | 92.6 | 7.4 | 100.0 | 127 779 | 11 075 | 138 854 | 92.0 | 8.0 | 100.0 |
|  | Female | 89 113 | 11 078 | 100 191 | 88.9 | 11.1 | 100.0 | 61 491 | 6 827 | 68 318 | 90.0 | 10.0 | 100.0 | 150 604 | 17 905 | 168 509 | 89.4 | 10.6 | 100.0 |
| Health  Condition | No | 154 357 | 15 508 | 169 865 | 90.9 | 9.1 | 100.0 | 80 324 | 6 914 | 87 238 | 92.1 | 7.9 | 100.0 | 234 681 | 22 422 | 257 103 | 91.3 | 8.7 | 100.0 |
|  | Yes | 20 309 | 3 263 | 23 572 | 86.2 | 13.8 | 100.0 | 23 393 | 3 295 | 26 688 | 87.7 | 12.4 | 100.0 | 43 702 | 6 558 | 50 260 | 87.0 | 13.0 | 100.0 |
| **Total** |  | **174 666** | **18 771** | **193 437** | **90.30** | **9.7** | **100** | **103 717** | **10 209** | **113 926** | **91.0** | **9.0** | **100.0** | **278 383** | **28 980** | **307 363** | **90.6** | **9.4** | **100.0** |

**b.**

|  |  | **Working** | | | | | | | | **Not working** | | | | | | | |
| --- | --- | --- | --- | --- | --- | --- | --- | --- | --- | --- | --- | --- | --- | --- | --- | --- | --- |
|  |  | **Count** | | | | **%** | | | | **Count** | | | | **%** | | | |
| **Outcome** | **Reduced function** | **No** | **Yes, a little** | **Yes, a lot** | **Total** | **No** | **Yes, a little** | **Yes, a lot** | **Total** | **No** | **Yes, a little** | **Yes, a lot** | **Total** | **No** | **Yes, a little** | **Yes, a lot** | **Total** |
| Age bands | 15-19 | 50 | 53 | 19 | 122 | 40.98 | 43.44 | 15.57 | 100 | 535 | 588 | 184 | 1 307 | 40.93 | 44.99 | 14.08 | 100 |
|  | 20-24 | 223 | 278 | 59 | 560 | 39.82 | 49.64 | 10.54 | 100 | 198 | 255 | 76 | 529 | 37.43 | 48.2 | 14.37 | 100 |
|  | 25-29 | 420 | 547 | 166 | 1133 | 37.07 | 48.28 | 14.65 | 100 | 95 | 129 | 75 | 299 | 31.77 | 43.14 | 25.08 | 100 |
|  | 30-34 | 484 | 778 | 276 | 1538 | 31.47 | 50.59 | 17.95 | 100 | 142 | 243 | 122 | 507 | 28.01 | 47.93 | 24.06 | 100 |
|  | 35-39 | 615 | 1 027 | 390 | 2032 | 30.27 | 50.54 | 19.19 | 100 | 173 | 301 | 209 | 683 | 25.33 | 44.07 | 30.6 | 100 |
|  | 40-44 | 835 | 1 368 | 551 | 2754 | 30.32 | 49.67 | 20.01 | 100 | 164 | 353 | 249 | 766 | 21.41 | 46.08 | 32.51 | 100 |
|  | 45-49 | 914 | 1 462 | 606 | 2982 | 30.65 | 49.03 | 20.32 | 100 | 171 | 369 | 284 | 824 | 20.75 | 44.78 | 34.47 | 100 |
|  | 50-54 | 870 | 1 547 | 741 | 3158 | 27.55 | 48.99 | 23.46 | 100 | 203 | 427 | 484 | 1 114 | 18.22 | 38.33 | 43.45 | 100 |
|  | 55-59 | 717 | 1 272 | 643 | 2632 | 27.24 | 48.33 | 24.43 | 100 | 283 | 663 | 569 | 1 515 | 18.68 | 43.76 | 37.56 | 100 |
|  | 60-64 | 423 | 810 | 433 | 1666 | 25.39 | 48.62 | 25.99 | 100 | 456 | 901 | 669 | 2 026 | 22.51 | 44.47 | 33.02 | 100 |
|  | 65-69 | 42 | 107 | 44 | 193 | 21.76 | 55.44 | 22.8 | 100 | 150 | 292 | 197 | 639 | 23.47 | 45.7 | 30.83 | 100 |
| Sex | Male | 2 493 | 3 818 | 1 381 | 7692 | 32.41 | 49.64 | 17.95 | 100 | 929 | 1 474 | 979 | 3 382 | 27.47 | 43.58 | 28.95 | 100 |
|  | Female | 3 100 | 5 431 | 2 547 | 11078 | 27.98 | 49.03 | 22.99 | 100 | 1 641 | 3 047 | 2 139 | 6 827 | 24.04 | 44.63 | 31.33 | 100 |
| Health  Condition | No | 4 895 | 7 711 | 2 902 | 15508 | 31.56 | 49.72 | 18.71 | 100 | 2 115 | 3 224 | 1 574 | 6 913 | 30.59 | 46.64 | 22.77 | 100 |
|  | Yes | 698 | 1 538 | 1 026 | 3262 | 29.80 | 49.28 | 31.45 | 100 | 455 | 1 297 | 1 544 | 3 296 | 13.80 | 39.35 | 46.84 | 100 |
| **Total** |  | **5 593** | **9 249** | **3 928** | **18 770** | **29.80** | **49.28** | **20.93** | **100** | **2 570** | **4 521** | **3 118** | **10 209** | **25.17** | **44.28** | **30.54** | **100** |

Table S7: OR for unadjusted and adjusted models for all 3 occupational exposure groups for the binary logistic model

| **Industry (SIC)** | | | | | | | **New occupations** | | | | | | |
| --- | --- | --- | --- | --- | --- | --- | --- | --- | --- | --- | --- | --- | --- |
|  | **Unadjusted** | | | **Adjusted** | | |  | **Unadjusted** | | | **Adjusted** | | |
|  |  | **CI95%** | |  | **CI95%** | |  |  | **CI95%** | |  | **CI95%** | |
| **Self-reported long-COVID** | **OR** | **Low** | **High** | **OR** | **Low** | **High** |  | **OR** | **Low** | **High** | **OR** | **Low** | **High** |
| Teaching and education | 1.41 | 1.36 | 1.46 | 1.27 | 1.23 | 1.31 | Education | 1.53 | 1.45 | 1.60 | 1.34 | 1.28 | 1.41 |
| Health care | 1.19 | 1.14 | 1.23 | 1.09 | 1.05 | 1.13 | Food processing | 1.16 | 0.97 | 1.38 | 1.10 | 0.92 | 1.31 |
| Social care | 1.38 | 1.28 | 1.48 | 1.22 | 1.13 | 1.31 | Healthcare-office based | 1.07 | 0.86 | 1.32 | 0.92 | 0.75 | 1.14 |
| Transport (incl. storage, logistic) | 1.08 | 1.00 | 1.16 | 1.12 | 1.04 | 1.21 | Healthcare-patient contact | 1.14 | 1.08 | 1.20 | 1.04 | 0.98 | 1.10 |
| Retail sector (incl. wholesale) | 1.08 | 1.03 | 1.14 | 1.07 | 1.02 | 1.13 | Hospitality | 1.27 | 1.13 | 1.43 | 1.27 | 1.13 | 1.43 |
| Hospitality (e.g. hotel, restaurant) | 1.01 | 0.92 | 1.12 | 1.05 | 0.95 | 1.16 | Manual | 0.94 | 0.89 | 1.00 | 1.03 | 0.97 | 1.10 |
| Food production, agriculture, farming | 0.96 | 0.86 | 1.07 | 0.99 | 0.89 | 1.11 | Other workers-non-office based | 0.93 | 0.88 | 0.98 | 0.98 | 0.93 | 1.05 |
| Personal services (e.g. hairdressers) | 0.98 | 0.85 | 1.14 | 0.94 | 0.81 | 1.10 | Other workers-office based | 0.90 | 0.88 | 0.92 | 0.90 | 0.88 | 0.92 |
| Information technology and communication | 0.75 | 0.71 | 0.79 | 0.81 | 0.77 | 0.86 | Personal care | 1.09 | 0.77 | 1.53 | 0.95 | 0.68 | 1.34 |
| Financial services incl. insurance | 0.78 | 0.74 | 0.82 | 0.81 | 0.77 | 0.85 | Police and protective services | 1.27 | 1.15 | 1.40 | 1.31 | 1.18 | 1.45 |
| Manufacturing or construction | 0.98 | 0.94 | 1.02 | 1.03 | 0.98 | 1.08 | Retail | 1.10 | 1.02 | 1.20 | 1.08 | 1.00 | 1.17 |
| Civil service or Local Government | 1.12 | 1.06 | 1.17 | 1.06 | 1.01 | 1.11 | Sanitation services | 1.18 | 1.02 | 1.36 | 1.03 | 0.89 | 1.19 |
| Armed forces | 0.77 | 0.60 | 0.99 | 0.86 | 0.67 | 1.11 | Social care | 1.53 | 1.44 | 1.63 | 1.34 | 1.26 | 1.43 |
| Arts,Entertainment or Recreation | 0.89 | 0.81 | 0.98 | 0.93 | 0.84 | 1.02 | Transport-nonpublic facing | 1.10 | 0.99 | 1.23 | 1.16 | 1.04 | 1.29 |
| Other occupation sector | 0.89 | 0.86 | 0.93 | 0.91 | 0.87 | 0.94 | Transport-public facing | 1.19 | 0.99 | 1.44 | 1.25 | 1.03 | 1.51 |
| Not working | 0.96 | 0.94 | 0.97 | 0.98 | 0.96 | 0.99 | Not working | 0.97 | 0.95 | 0.98 | 0.99 | 0.97 | 1.00 |
|  |  |  |  |  |  |  | **Occupation (SOC-1 digit)** | | | | | | |
|  |  |  |  |  |  |  |  | **Unadjusted** | | | **Adjusted** | | |
|  |  |  |  |  |  |  |  |  | **CI95%** | |  | **CI95%** | |
|  |  |  |  |  |  |  |  | **OR** | **Low** | **High** | **OR** | **Low** | **High** |
|  |  |  |  |  |  |  | Managers, directors and senior official | 0.99 | 0.95 | 1.03 | 0.99 | 0.95 | 1.03 |
|  |  |  |  |  |  |  | Professional occupations | 0.97 | 0.95 | 0.99 | 0.96 | 0.94 | 0.99 |
|  |  |  |  |  |  |  | Associate professional and technical | 0.98 | 0.95 | 1.02 | 1.01 | 0.97 | 1.04 |
|  |  |  |  |  |  |  | Admin and secretarial | 0.99 | 0.95 | 1.03 | 0.91 | 0.87 | 0.95 |
|  |  |  |  |  |  |  | Skilled trades | 0.94 | 0.89 | 1.00 | 1.04 | 0.98 | 1.11 |
|  |  |  |  |  |  |  | Caring, leisure and other service | 1.65 | 1.58 | 1.73 | 1.44 | 1.38 | 1.52 |
|  |  |  |  |  |  |  | Sales and customer service | 1.16 | 1.08 | 1.24 | 1.11 | 1.04 | 1.19 |
|  |  |  |  |  |  |  | Process plant and machine operatives | 1.07 | 0.99 | 1.16 | 1.14 | 1.05 | 1.23 |
|  |  |  |  |  |  |  | Elementary | 1.12 | 1.05 | 1.20 | 1.11 | 1.03 | 1.19 |
|  |  |  |  |  |  |  | Not working | 0.95 | 0.94 | 0.97 | 0.97 | 0.95 | 0.99 |

Table S8: OR for unadjusted and adjusted models for all 3 occupational exposure groups for the ordered logistic model

| **Industry (SIC)** | | | | | | | **New occupations** | | | | | | |
| --- | --- | --- | --- | --- | --- | --- | --- | --- | --- | --- | --- | --- | --- |
|  | **Unadjusted** | | | **Adjusted** | | |  | **Unadjusted** | | | **Adjusted** | | |
|  |  | **CI95%** | |  | **CI95%** | |  |  | **CI95%** | |  | **CI95%** | |
| **Self-reported long-COVID** | **OR** | **Low** | **High** | **OR** | **Low** | **High** |  | **OR** | **Low** | **High** | **OR** | **Low** | **High** |
| Teaching and education | 0.98 | 0.93 | 1.03 | 0.98 | 0.92 | 1.03 | Education | 0.97 | 0.90 | 1.05 | 0.97 | 0.89 | 1.05 |
| Health care | 1.07 | 1.00 | 1.14 | 1.00 | 0.93 | 1.07 | Food processing | 0.95 | 0.69 | 1.31 | 0.86 | 0.62 | 1.21 |
| Social care | 1.07 | 0.95 | 1.20 | 0.96 | 0.85 | 1.08 | Healthcare-office based | 0.79 | 0.52 | 1.19 | 0.70 | 0.46 | 1.08 |
| Transport (incl. storage, logistic) | 0.90 | 0.79 | 1.02 | 0.94 | 0.82 | 1.08 | Healthcare-patient contact | 1.03 | 0.94 | 1.13 | 0.98 | 0.89 | 1.08 |
| Retail sector (incl. wholesale) | 0.89 | 0.81 | 0.98 | 0.91 | 0.83 | 1.00 | Hospitality | 0.89 | 0.72 | 1.09 | 0.91 | 0.74 | 1.12 |
| Hospitality (e.g. hotel, restaurant) | 0.76 | 0.64 | 0.90 | 0.82 | 0.68 | 0.98 | Manual | 0.91 | 0.82 | 1.02 | 1.01 | 0.90 | 1.13 |
| Food production, agriculture, farming | 0.99 | 0.82 | 1.20 | 0.98 | 0.80 | 1.19 | Other workers-non-office based | 0.74 | 0.67 | 0.82 | 0.82 | 0.74 | 0.91 |
| Personal services (e.g. hairdressers) | 0.95 | 0.73 | 1.23 | 0.94 | 0.71 | 1.23 | Other workers-office based | 0.76 | 0.73 | 0.79 | 0.79 | 0.76 | 0.82 |
| Information technology and communication | 0.69 | 0.63 | 0.77 | 0.79 | 0.71 | 0.88 | Personal care | 1.25 | 0.67 | 2.32 | 1.21 | 0.64 | 2.27 |
| Financial services incl. insurance | 0.69 | 0.63 | 0.75 | 0.76 | 0.69 | 0.83 | Police and protective services | 0.90 | 0.75 | 1.07 | 0.96 | 0.80 | 1.16 |
| Manufacturing or construction | 0.79 | 0.73 | 0.85 | 0.87 | 0.80 | 0.94 | Retail | 1.00 | 0.87 | 1.14 | 1.04 | 0.91 | 1.19 |
| Civil service or Local Government | 0.99 | 0.91 | 1.08 | 0.94 | 0.86 | 1.03 | Sanitation services | 0.94 | 0.74 | 1.21 | 0.80 | 0.62 | 1.03 |
| Armed forces | 0.94 | 0.59 | 1.50 | 1.18 | 0.72 | 1.94 | Social care | 1.23 | 1.10 | 1.37 | 1.09 | 0.97 | 1.22 |
| Arts,Entertainment or Recreation | 0.73 | 0.62 | 0.87 | 0.81 | 0.68 | 0.97 | Transport-nonpublic facing | 0.81 | 0.67 | 0.97 | 0.83 | 0.69 | 1.00 |
| Other occupation sector | 0.81 | 0.76 | 0.87 | 0.85 | 0.80 | 0.91 | Transport-public facing | 1.29 | 0.92 | 1.81 | 1.28 | 0.91 | 1.80 |
| Not working | 1.26 | 1.23 | 1.30 | 1.21 | 1.17 | 1.26 | Not working | 1.25 | 1.21 | 1.29 | 1.21 | 1.18 | 1.25 |
|  |  |  |  |  |  |  | **Occupation (SOC-1 digit)** | | | | | | |
|  |  |  |  |  |  |  |  | **Unadjusted** | | | **Adjusted** | | |
|  |  |  |  |  |  |  |  |  | **CI95%** | |  | **CI95%** | |
|  |  |  |  |  |  |  |  | **OR** | **Low** | **High** | **OR** | **Low** | **High** |
|  |  |  |  |  |  |  | Managers, directors and senior official | 0.77 | 0.71 | 0.83 | 0.82 | 0.75 | 0.88 |
|  |  |  |  |  |  |  | Professional occupations | 0.88 | 0.84 | 0.92 | 0.92 | 0.88 | 0.96 |
|  |  |  |  |  |  |  | Associate professional and technical | 0.81 | 0.77 | 0.86 | 0.87 | 0.82 | 0.92 |
|  |  |  |  |  |  |  | Admin and secretarial | 0.87 | 0.81 | 0.93 | 0.83 | 0.77 | 0.89 |
|  |  |  |  |  |  |  | Skilled trades | 0.83 | 0.75 | 0.92 | 0.93 | 0.84 | 1.04 |
|  |  |  |  |  |  |  | Caring, leisure and other service | 1.09 | 1.01 | 1.18 | 1.01 | 0.93 | 1.10 |
|  |  |  |  |  |  |  | Sales and customer service | 0.93 | 0.83 | 1.05 | 0.92 | 0.81 | 1.03 |
|  |  |  |  |  |  |  | Process plant and machine operatives | 0.97 | 0.85 | 1.12 | 1.01 | 0.87 | 1.16 |
|  |  |  |  |  |  |  | Elementary | 0.96 | 0.84 | 1.08 | 0.92 | 0.81 | 1.04 |
|  |  |  |  |  |  |  | Not working | 1.27 | 1.23 | 1.31 | 1.22 | 1.18 | 1.26 |

Table S9: Probabilities (margins) of self-reported long-COVID and reduced function because of COVID by industry (SIC) occupations and major occupational SOC groups (95% CIs)

|  |  | **Self-reported long-COVID** | | | **Reduced function** | | | | | | | | |
| --- | --- | --- | --- | --- | --- | --- | --- | --- | --- | --- | --- | --- | --- |
|  |  | **Yes** | | | **No** | | | **Yes, a little** | | | **Yes, a lot** | | |
|  |  |  | **CI95%** | |  | **CI95%** | |  | **CI95%** | |  | **CI95%** | |
|  | **Self-reported long-COVID** | **Prob** | **Low** | **High** | **Prob** | **Low** | **High** | **Prob** | **Low** | **High** | **Prob** | **Low** | **High** |
| Industry (SIC) | Teaching and education | 0.116 | 0.112 | 0.119 | 0.290 | 0.276 | 0.305 | 0.493 | 0.485 | 0.501 | 0.217 | 0.205 | 0.228 |
|  | Health care | 0.101 | 0.097 | 0.105 | 0.301 | 0.282 | 0.319 | 0.491 | 0.483 | 0.499 | 0.208 | 0.193 | 0.223 |
|  | Social care | 0.111 | 0.104 | 0.118 | 0.319 | 0.288 | 0.351 | 0.486 | 0.475 | 0.498 | 0.194 | 0.172 | 0.217 |
|  | Transport (incl. storage, logistic) | 0.104 | 0.097 | 0.110 | 0.313 | 0.280 | 0.347 | 0.488 | 0.477 | 0.499 | 0.199 | 0.174 | 0.224 |
|  | Retail sector (incl. wholesale) | 0.100 | 0.095 | 0.104 | 0.313 | 0.290 | 0.336 | 0.488 | 0.479 | 0.498 | 0.199 | 0.182 | 0.217 |
|  | Hospitality (e.g. hotel, restaurant) | 0.097 | 0.089 | 0.106 | 0.304 | 0.271 | 0.337 | 0.490 | 0.480 | 0.501 | 0.206 | 0.180 | 0.231 |
|  | Food production, agriculture, farming | 0.093 | 0.084 | 0.102 | 0.306 | 0.254 | 0.359 | 0.490 | 0.475 | 0.504 | 0.204 | 0.164 | 0.244 |
|  | Personal services (e.g. hairdressers) | 0.089 | 0.077 | 0.101 | 0.302 | 0.250 | 0.353 | 0.491 | 0.477 | 0.504 | 0.208 | 0.167 | 0.248 |
|  | Information tech & communication | 0.078 | 0.073 | 0.082 | 0.334 | 0.308 | 0.359 | 0.482 | 0.471 | 0.493 | 0.184 | 0.166 | 0.202 |
|  | Financial services incl. insurance | 0.077 | 0.073 | 0.081 | 0.345 | 0.320 | 0.370 | 0.478 | 0.467 | 0.489 | 0.177 | 0.160 | 0.193 |
|  | Manufacturing or construction | 0.096 | 0.092 | 0.100 | 0.323 | 0.303 | 0.344 | 0.485 | 0.476 | 0.494 | 0.191 | 0.176 | 0.206 |
|  | Civil service or Local Government | 0.098 | 0.094 | 0.103 | 0.318 | 0.295 | 0.341 | 0.487 | 0.477 | 0.496 | 0.195 | 0.178 | 0.212 |
|  | Armed forces | 0.082 | 0.063 | 0.100 | 0.270 | 0.160 | 0.381 | 0.496 | 0.483 | 0.509 | 0.234 | 0.134 | 0.333 |
|  | Arts, Entertainment or Recreation | 0.087 | 0.079 | 0.095 | 0.355 | 0.309 | 0.401 | 0.475 | 0.456 | 0.493 | 0.171 | 0.142 | 0.200 |
|  | Other occupation sector | 0.086 | 0.083 | 0.089 | 0.327 | 0.310 | 0.344 | 0.484 | 0.475 | 0.493 | 0.189 | 0.176 | 0.201 |
|  | Not working | 0.092 | 0.090 | 0.093 | 0.231 | 0.217 | 0.245 | 0.496 | 0.489 | 0.504 | 0.273 | 0.258 | 0.287 |
| New occupations | Education | 0.120 | 0.115 | 0.125 | 0.282 | 0.266 | 0.298 | 0.477 | 0.471 | 0.484 | 0.241 | 0.226 | 0.255 |
|  | Food processing | 0.101 | 0.085 | 0.117 | 0.304 | 0.236 | 0.373 | 0.474 | 0.460 | 0.488 | 0.221 | 0.166 | 0.277 |
|  | Healthcare-office based | 0.086 | 0.070 | 0.103 | 0.347 | 0.255 | 0.440 | 0.463 | 0.434 | 0.493 | 0.189 | 0.126 | 0.253 |
|  | Healthcare-patient contact | 0.096 | 0.091 | 0.101 | 0.280 | 0.261 | 0.300 | 0.477 | 0.471 | 0.484 | 0.242 | 0.225 | 0.260 |
|  | Hospitality | 0.114 | 0.102 | 0.126 | 0.294 | 0.253 | 0.335 | 0.476 | 0.467 | 0.484 | 0.230 | 0.195 | 0.266 |
|  | Manual | 0.095 | 0.090 | 0.101 | 0.273 | 0.252 | 0.295 | 0.478 | 0.472 | 0.484 | 0.249 | 0.228 | 0.269 |
|  | Other workers-non-office based | 0.091 | 0.086 | 0.096 | 0.316 | 0.294 | 0.338 | 0.472 | 0.464 | 0.479 | 0.212 | 0.195 | 0.230 |
|  | Other workers-office based | 0.084 | 0.082 | 0.086 | 0.323 | 0.313 | 0.333 | 0.470 | 0.464 | 0.476 | 0.207 | 0.199 | 0.215 |
|  | Personal care | 0.089 | 0.061 | 0.116 | 0.241 | 0.130 | 0.352 | 0.478 | 0.465 | 0.490 | 0.281 | 0.160 | 0.403 |
|  | Police and protective services | 0.118 | 0.107 | 0.128 | 0.284 | 0.247 | 0.320 | 0.477 | 0.470 | 0.484 | 0.239 | 0.206 | 0.272 |
|  | Retail | 0.099 | 0.092 | 0.106 | 0.268 | 0.242 | 0.294 | 0.478 | 0.472 | 0.484 | 0.254 | 0.228 | 0.279 |
|  | Sanitation services | 0.095 | 0.083 | 0.107 | 0.320 | 0.267 | 0.374 | 0.471 | 0.457 | 0.485 | 0.209 | 0.168 | 0.250 |
|  | Social care | 0.120 | 0.113 | 0.127 | 0.260 | 0.239 | 0.281 | 0.478 | 0.472 | 0.485 | 0.262 | 0.241 | 0.283 |
|  | Transport-nonpublic facing | 0.105 | 0.095 | 0.116 | 0.313 | 0.273 | 0.352 | 0.472 | 0.462 | 0.483 | 0.215 | 0.184 | 0.246 |
|  | Transport-public facing | 0.113 | 0.094 | 0.132 | 0.231 | 0.172 | 0.289 | 0.476 | 0.465 | 0.487 | 0.293 | 0.226 | 0.360 |
|  | Not working | 0.092 | 0.090 | 0.093 | 0.240 | 0.232 | 0.248 | 0.477 | 0.471 | 0.484 | 0.282 | 0.274 | 0.291 |
| Occupation (SOC-1 digit) | Managers, directors and senior official | 0.093 | 0.089 | 0.097 | 0.320 | 0.303 | 0.337 | 0.472 | 0.465 | 0.478 | 0.209 | 0.195 | 0.222 |
|  | Professional occupations | 0.091 | 0.088 | 0.093 | 0.296 | 0.286 | 0.306 | 0.476 | 0.470 | 0.482 | 0.228 | 0.219 | 0.237 |
|  | Associate professional and technical | 0.094 | 0.091 | 0.098 | 0.307 | 0.294 | 0.321 | 0.474 | 0.468 | 0.480 | 0.219 | 0.207 | 0.230 |
|  | Admin and secretarial | 0.086 | 0.083 | 0.090 | 0.317 | 0.301 | 0.333 | 0.472 | 0.465 | 0.479 | 0.211 | 0.198 | 0.223 |
|  | Skilled trades | 0.097 | 0.092 | 0.103 | 0.293 | 0.271 | 0.315 | 0.477 | 0.470 | 0.483 | 0.230 | 0.212 | 0.249 |
|  | Caring, leisure and other service | 0.130 | 0.124 | 0.135 | 0.277 | 0.260 | 0.294 | 0.478 | 0.473 | 0.484 | 0.244 | 0.229 | 0.260 |
|  | Sales and customer service | 0.103 | 0.097 | 0.109 | 0.296 | 0.271 | 0.320 | 0.476 | 0.469 | 0.483 | 0.228 | 0.207 | 0.249 |
|  | Process plant and machine operatives | 0.105 | 0.097 | 0.113 | 0.278 | 0.249 | 0.306 | 0.478 | 0.472 | 0.485 | 0.244 | 0.218 | 0.270 |
|  | Elementary | 0.103 | 0.096 | 0.109 | 0.295 | 0.270 | 0.321 | 0.476 | 0.469 | 0.483 | 0.228 | 0.206 | 0.250 |
|  | Not working | 0.091 | 0.090 | 0.093 | 0.242 | 0.234 | 0.250 | 0.478 | 0.473 | 0.484 | 0.280 | 0.271 | 0.288 |

Table S10: Comparison of odds ratios between one observation person and Panel analyses (Industrial SIC groups)

| **Industries** | **Long Covid (adjusted model)** | | | | | | **Reduced function (adjusted model)** | | | | | |
| --- | --- | --- | --- | --- | --- | --- | --- | --- | --- | --- | --- | --- |
|  | **One observation per person** | | | **Panel** | | | **One observation per person** | | | **Panel** | | |
|  | **OR** | **95% CI** | | **OR** | **95% CI** | | **OR** | **95% CI** | | **OR** | **95% CI** | |
|  |  | **Lower** | **Higher** |  | **Lower** | **Higher** |  | **Lower** | **Higher** |  | **Lower** | **Higher** |
| **Teaching and education** | 1.27 | 1.23 | 1.31 | 1.39 | 1.31 | 1.49 | 0.98 | 0.92 | 1.03 | 0.92 | 0.85 | 0.98 |
| **Health care** | 1.09 | 1.05 | 1.13 | 1.25 | 1.17 | 1.35 | 1.00 | 0.93 | 1.07 | 0.85 | 0.79 | 0.92 |
| **Social care** | 1.22 | 1.13 | 1.31 | 1.28 | 1.12 | 1.45 | 0.96 | 0.85 | 1.08 | 0.82 | 0.71 | 0.94 |
| **Transport (incl. storage, logistic)** | 1.12 | 1.04 | 1.21 | 1.20 | 1.03 | 1.40 | 0.94 | 0.82 | 1.08 | 0.91 | 0.78 | 1.07 |
| **Retail sector (incl. wholesale)** | 1.07 | 1.02 | 1.13 | 1.05 | 0.95 | 1.16 | 0.91 | 0.83 | 1.00 | 0.87 | 0.78 | 0.97 |
| **Hospitality (e.g. hotel, restaurant)** | 1.05 | 0.95 | 1.16 | 1.15 | 1.00 | 1.32 | 0.82 | 0.68 | 0.98 | 0.89 | 0.75 | 1.06 |
| **Food production, agriculture, farming** | 0.99 | 0.89 | 1.11 | 0.94 | 0.73 | 1.21 | 0.98 | 0.80 | 1.19 | 1.03 | 0.83 | 1.29 |
| **Personal services (e.g. hairdressers)** | 0.94 | 0.81 | 1.10 | 0.93 | 0.74 | 1.16 | 0.94 | 0.71 | 1.23 | 0.88 | 0.68 | 1.14 |
| **Information technology and communication** | 0.81 | 0.77 | 0.86 | 0.69 | 0.59 | 0.81 | 0.79 | 0.71 | 0.88 | 0.70 | 0.62 | 0.80 |
| **Financial services incl. insurance** | 0.81 | 0.77 | 0.85 | 0.68 | 0.59 | 0.79 | 0.76 | 0.69 | 0.83 | 0.68 | 0.60 | 0.76 |
| **Manufacturing or construction** | 1.03 | 0.98 | 1.08 | 1.08 | 0.98 | 1.20 | 0.87 | 0.80 | 0.94 | 0.82 | 0.74 | 0.90 |
| **Civil service or Local Government** | 1.06 | 1.01 | 1.11 | 1.10 | 1.00 | 1.21 | 0.94 | 0.86 | 1.03 | 0.84 | 0.76 | 0.93 |
| **Armed forces** | 0.86 | 0.67 | 1.11 | 0.73 | 0.35 | 1.52 | 1.18 | 0.72 | 1.94 | 1.38 | 0.73 | 2.63 |
| **Arts,Entertainment or Recreation** | 0.93 | 0.84 | 1.02 | 0.90 | 0.73 | 1.10 | 0.81 | 0.68 | 0.97 | 0.85 | 0.69 | 1.04 |
| **Other occupation sector** | 0.91 | 0.87 | 0.94 | 0.89 | 0.82 | 0.96 | 0.85 | 0.80 | 0.91 | 0.75 | 0.70 | 0.82 |
| **Not working** | 0.98 | 0.96 | 0.99 | 0.95 | 0.92 | 0.98 | 1.21 | 1.17 | 1.26 | 1.51 | 1.44 | 1.57 |

Table S11: Comparison of odds ratios between one observation person and Panel analyses (Occupational groups)

| **Occupations** | **Long Covid (adjusted model)** | | | | | | **Reduced function (adjusted model)** | | | | | |
| --- | --- | --- | --- | --- | --- | --- | --- | --- | --- | --- | --- | --- |
|  | **One observation per person** | | | **Panel** | | | **One observation per person** | | | **Panel** | | |
|  | **OR** | **95% CI** | | **OR** | **95% CI** | | **OR** | **95% CI** | | **OR** | **95% CI** | |
|  |  | **Lower** | **Higher** |  | **Lower** | **Higher** |  | **Lower** | **Higher** |  | **Lower** | **Higher** |
| **Education** | 1.34 | 1.28 | 1.41 | 1.58 | 1.45 | 1.72 | 0.97 | 0.89 | 1.05 | 0.90 | 0.82 | 1.00 |
| **food processing** | 1.10 | 0.92 | 1.31 | 0.99 | 0.75 | 1.32 | 0.86 | 0.62 | 1.21 | 0.82 | 0.58 | 1.16 |
| **Healthcare-office based** | 0.92 | 0.75 | 1.14 | 0.80 | 0.52 | 1.25 | 0.70 | 0.46 | 1.08 | 0.61 | 0.37 | 1.00 |
| **Healthcare-patient contact** | 1.04 | 0.98 | 1.10 | 1.17 | 1.05 | 1.30 | 0.98 | 0.89 | 1.08 | 0.87 | 0.78 | 0.98 |
| **Hospitality** | 1.27 | 1.13 | 1.43 | 1.37 | 1.16 | 1.63 | 0.91 | 0.74 | 1.12 | 0.94 | 0.76 | 1.16 |
| **Manual** | 1.03 | 0.97 | 1.10 | 1.08 | 0.94 | 1.24 | 1.01 | 0.90 | 1.13 | 0.94 | 0.83 | 1.08 |
| **Other workers-non-office based** | 0.98 | 0.93 | 1.05 | 0.99 | 0.87 | 1.13 | 0.82 | 0.74 | 0.91 | 0.81 | 0.72 | 0.93 |
| **Other workers-office based** | 0.90 | 0.88 | 0.92 | 0.86 | 0.82 | 0.90 | 0.79 | 0.76 | 0.82 | 0.69 | 0.66 | 0.72 |
| **Personal care** | 0.95 | 0.68 | 1.34 | 1.16 | 0.85 | 1.60 | 1.21 | 0.64 | 2.27 | 0.77 | 0.53 | 1.13 |
| **Police and protective services** | 1.31 | 1.18 | 1.45 | 1.69 | 1.38 | 2.07 | 0.96 | 0.80 | 1.16 | 0.83 | 0.67 | 1.03 |
| **Retail** | 1.08 | 1.00 | 1.17 | 1.18 | 1.02 | 1.35 | 1.04 | 0.91 | 1.19 | 0.96 | 0.83 | 1.11 |
| **Sanitation services** | 1.03 | 0.89 | 1.19 | 0.98 | 0.76 | 1.27 | 0.80 | 0.62 | 1.03 | 0.70 | 0.53 | 0.93 |
| **Social care** | 1.34 | 1.26 | 1.43 | 1.44 | 1.29 | 1.62 | 1.09 | 0.97 | 1.22 | 0.83 | 0.73 | 0.94 |
| **Transport-nonpublic facing** | 1.16 | 1.04 | 1.29 | 1.12 | 0.88 | 1.43 | 0.83 | 0.69 | 1.00 | 0.67 | 0.53 | 0.85 |
| **Transport-public facing** | 1.25 | 1.03 | 1.51 | 1.43 | 1.04 | 1.95 | 1.28 | 0.91 | 1.80 | 0.91 | 0.65 | 1.27 |
| **Not working** | 0.99 | 0.97 | 1.00 | 0.95 | 0.92 | 0.98 | 1.21 | 1.18 | 1.25 | 1.52 | 1.45 | 1.58 |

Table S12: Comparison of odds ratios between one observation person and Panel analyses (1-digit SOC groups)

| **1-digit SOC** | **Long Covid (adjusted model)** | | | | | | **Reduced function (adjusted model)** | | | | | |
| --- | --- | --- | --- | --- | --- | --- | --- | --- | --- | --- | --- | --- |
|  | **One observation per person** | | | **Panel** | | | **One observation per person** | | | **Panel** | | |
|  | **OR** | **95% CI** | | **OR** | **95% CI** | | **OR** | **95% CI** | | **OR** | **95% CI** | |
|  |  | **Lower** | **Higher** |  | **Lower** | **Higher** |  | **Lower** | **Higher** |  | **Lower** | **Higher** |
| **Managers, directors and senior official** | 0.99 | 0.95 | 1.03 | 0.94 | 0.90 | 1.09 | 0.82 | 0.75 | 0.88 | 0.73 | 0.66 | 0.81 |
| **Professional occupations** | 0.96 | 0.94 | 0.99 | 1.03 | 0.89 | 0.99 | 0.92 | 0.88 | 0.96 | 0.84 | 0.80 | 0.89 |
| **Associate professional and technical** | 1.01 | 0.97 | 1.04 | 0.88 | 0.96 | 1.11 | 0.87 | 0.82 | 0.92 | 0.78 | 0.72 | 0.84 |
| **Admin and secretarial** | 0.91 | 0.87 | 0.95 | 1.15 | 0.81 | 0.95 | 0.83 | 0.77 | 0.89 | 0.71 | 0.65 | 0.77 |
| **Skilled trades** | 1.04 | 0.98 | 1.11 | 1.56 | 1.01 | 1.31 | 0.93 | 0.84 | 1.04 | 0.92 | 0.81 | 1.05 |
| **Caring, leisure and other service** | 1.44 | 1.38 | 1.52 | 1.11 | 1.44 | 1.69 | 1.01 | 0.93 | 1.10 | 0.86 | 0.78 | 0.94 |
| **Sales and customer service** | 1.11 | 1.04 | 1.19 | 1.30 | 0.99 | 1.24 | 0.92 | 0.81 | 1.03 | 0.91 | 0.80 | 1.05 |
| **Process plant and machine operatives** | 1.14 | 1.05 | 1.23 | 1.09 | 1.11 | 1.52 | 1.01 | 0.87 | 1.16 | 0.85 | 0.72 | 1.01 |
| **Elementary** | 1.11 | 1.03 | 1.19 | 0.95 | 0.97 | 1.23 | 0.92 | 0.81 | 1.04 | 0.89 | 0.77 | 1.02 |
| **Not working** | 0.97 | 0.95 | 0.99 | 0.94 | 0.91 | 0.98 | 1.22 | 1.18 | 1.26 | 1.52 | 1.46 | 1.59 |

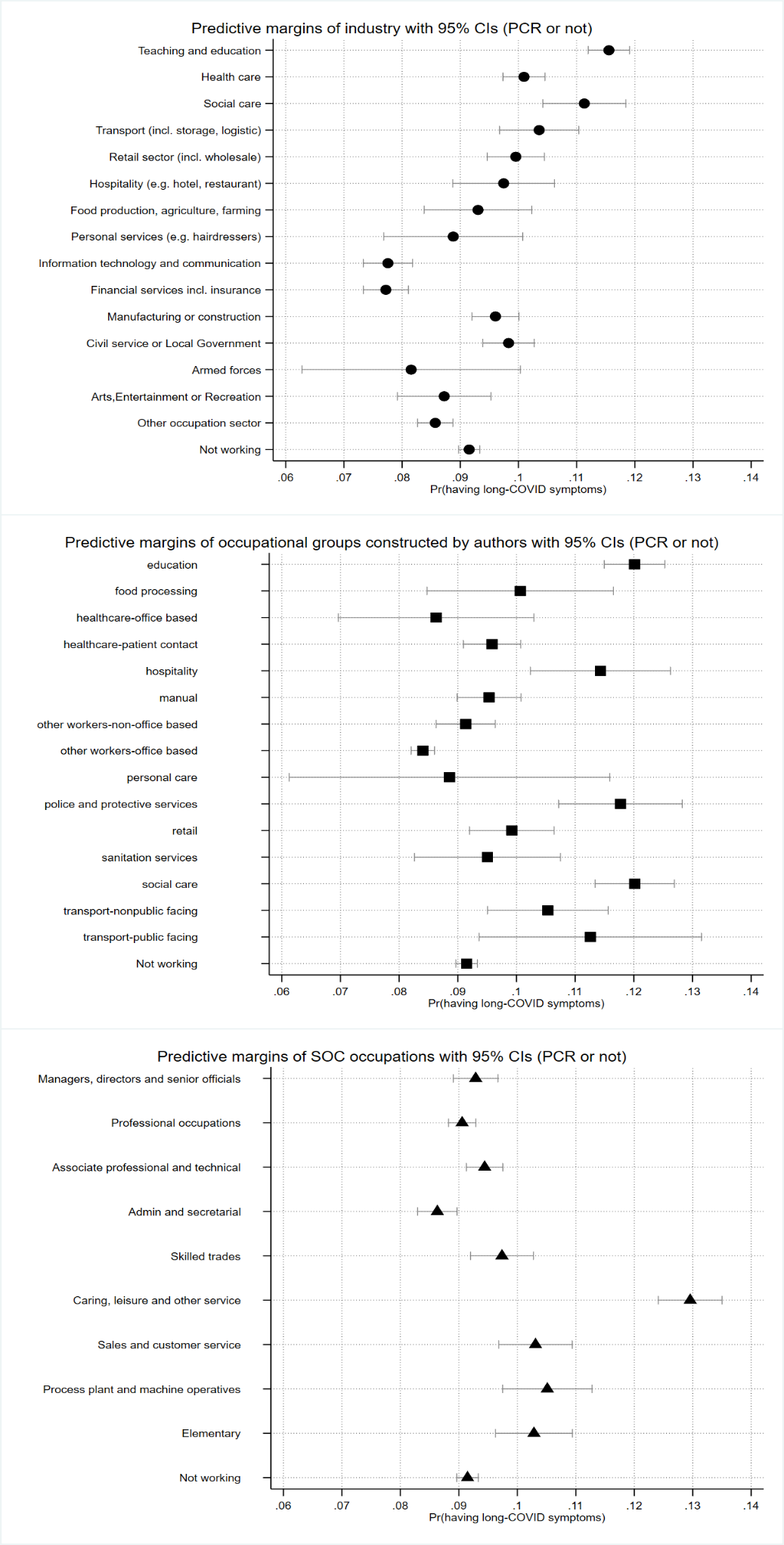

**c.**

**b.**

**a.**

Figure S1: Predictive margins of long-COVID by (a) industry; (b) occupational groups; and (c) major SOC groups

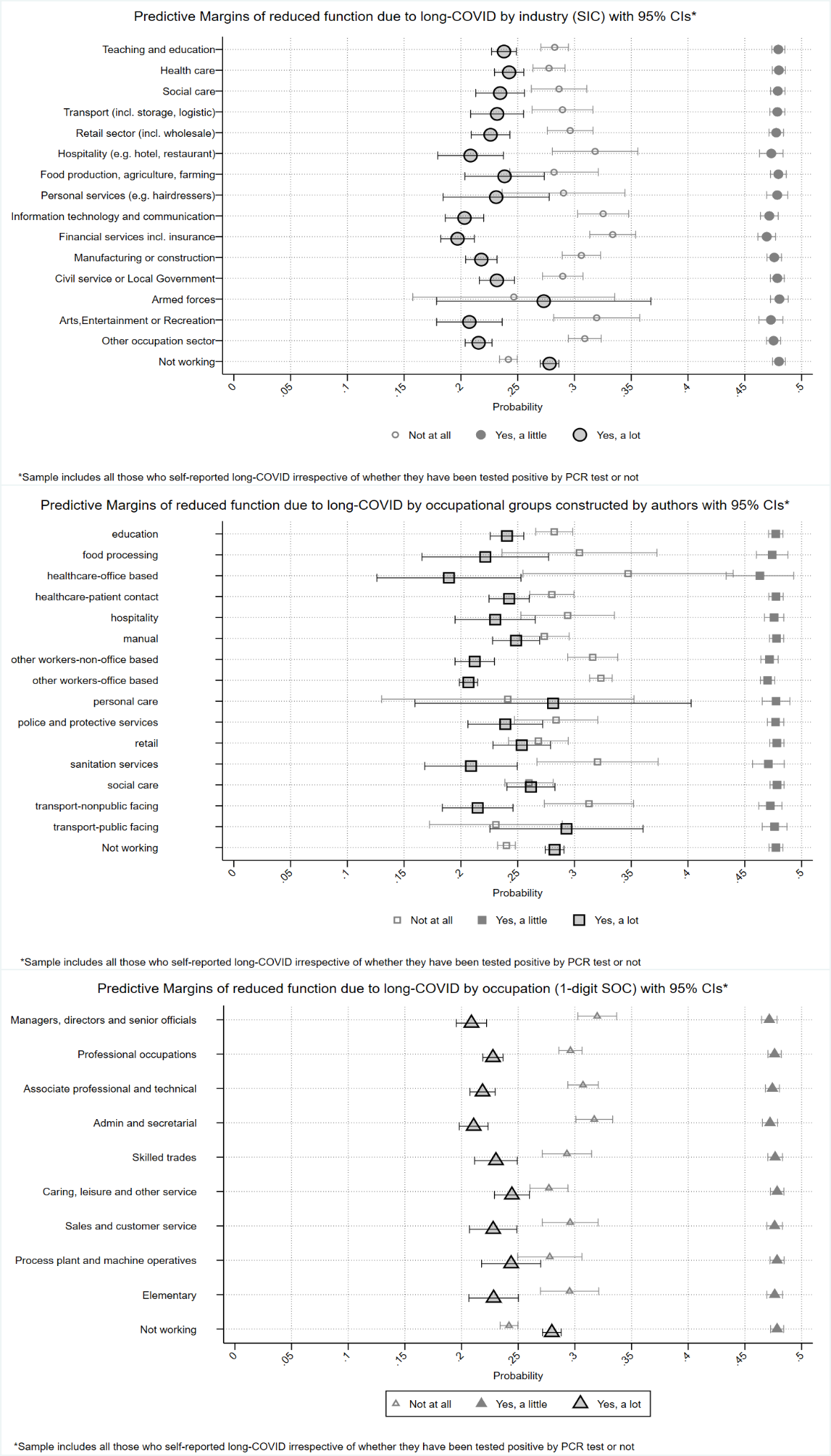

**b.**

**c.**

**a.**

Figure S2: Predictive margins of reduced activities due to long-COVID by (a) industry; (b) occupational groups; and (c) major SOC groups

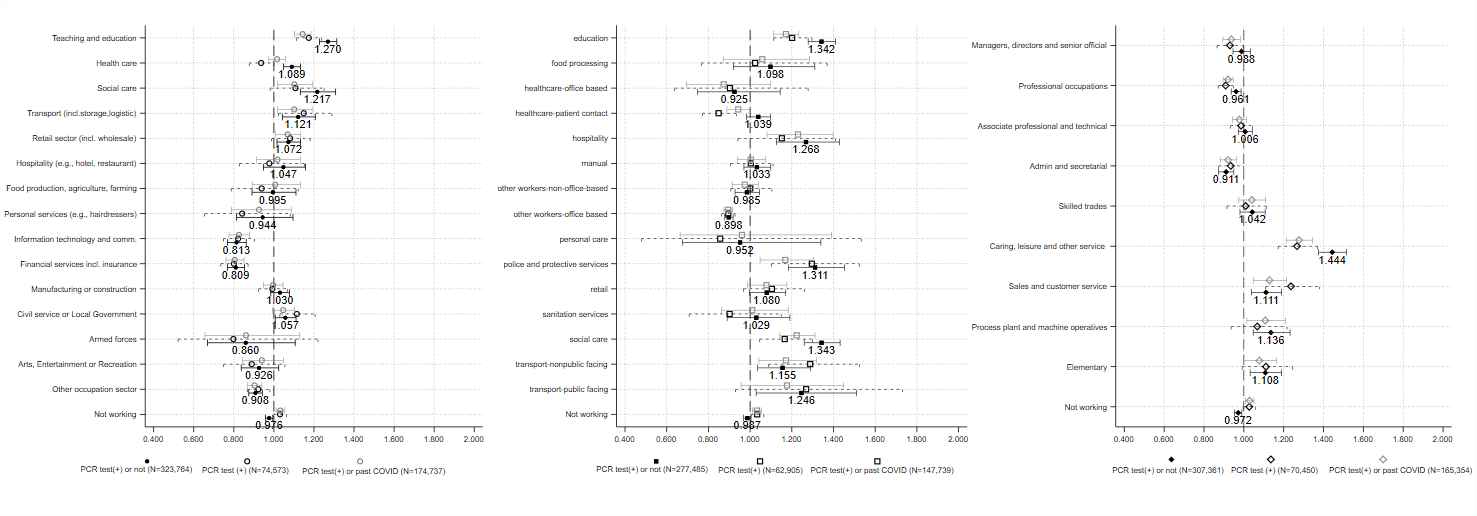

Figure S3: OR comparison between all three samples across all exposure groups (i: PCR test(+) or not, ii: PCR test (+) and iii) PCR test (+) or past COVID) (Long-COVID)

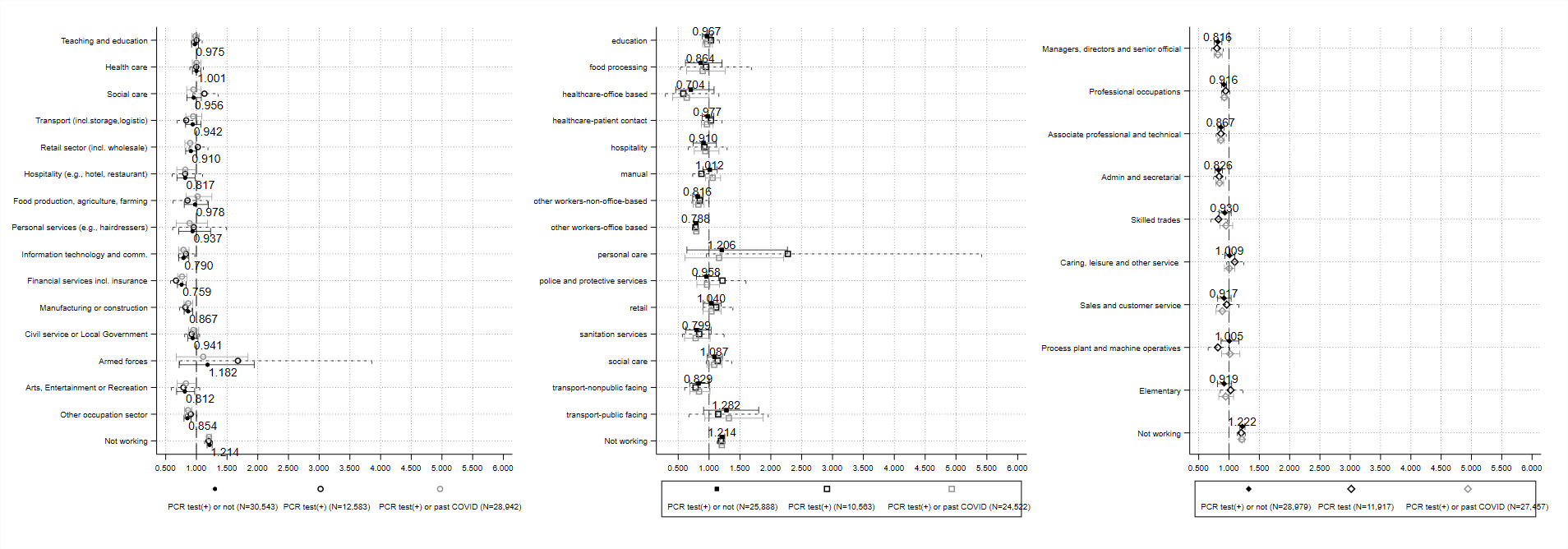

FigureS4: OR comparison between all three samples across all exposure groups (i: PCR test(+) or not, ii: PCR test (+) and iii) PCR test (+) or past COVID) (Reduced function)
